# A Functional *FCGR2A* Haplotype Associated with Rheumatoid Arthritis Determines Circulating Soluble FcγRIIa Levels and Immune Complex Signalling

**DOI:** 10.64898/2026.08.16.26360492

**Authors:** Euan W. Baxter, Eleanor G. Foy, John C. Taylor, Maren Thomsen, Sina Bondza, Simon E. Kolstoe, Steve Eyre, BRAGGSS Consortium, YEAR, Anthony G. Wilson, John D Isaacs, Paul Emery, Javier Martin, Mattia Frontini, Tolulope Balogun, NIHR BioResource – Rare Diseases RNA Consortium, Anne L. Barton, Adrian Goldman, Jennifer H. Barrett, Ann W. Morgan, James I. Robinson

**Affiliations:** Discovery and Translational Sciences Department, Leeds Institute of Cardiovascular and Metabolic Medicine, University of Leeds, UK; NIHR Leeds Biomedical Research Centre, Leeds Teaching Hospitals NHS Trust, UK; Leeds Institute of Medical Research, University of Leeds, UK; Astbury Centre for Structural and Molecular Biology, University of Leeds, Leeds, UK; Department of Immunology, Genetics and Pathology, Uppsala University, Uppsala, Sweden; Ridgeview Instruments AB, Uppsala, Sweden; School of Health and Care Professions, University of Portsmouth, Portsmouth, UK; Centre for Musculoskeletal Research, Manchester Academic Health Science Centre, The University of Manchester, Manchester, UK and NIHR Manchester Biomedical Research Centre, Manchester University NHS Trust, Manchester, UK; Biologics in Rheumatoid Arthritis Genetics and Genomics Syndicate, UK; Yorkshire Early Arthritis Register (YEAR); UCD School of Medicine and Medical Science, Conway Institute, University College Dublin, Dublin, Ireland; Musculoskeletal Research Group, Translational and Clinical Research Institute, Newcastle University and NIHR Newcastle Biomedical Research Centre, Newcastle upon Tyne Hospitals NHS Foundation Trust, Newcastle upon Tyne, UK; Instituto de Parasitologia y Biomedicina Lopez-Neyra, CSIC, Granada, Spain; Dept of Clinical and Biomedical Sciences, University of Exeter, Exeter, UK; NIHR BioResource – Cambridge Biomedical Campus, Cambridge, CB2 0QQ, UK; Faculty of Biological and Environmental Sciences, University of Helsinki, 00100 Helsinki, Finland

**Keywords:** Fc gamma receptor IIa, FcγRIIa, Q27W, H131R, IgG, immune complexes

## Abstract

FcγRIIa, encoded by *FCGR2A*, is a widely expressed Fc receptor implicated in autoimmunity and infectious disease susceptibility. To fine-map the rheumatoid arthritis (RA) association at the complex *FCGR* locus, we combined gene-specific resequencing, genetic association studies in UK and Spanish European cohorts, functional genomics, structural biology, biophysical analyses, and cellular assays. We identified a common European *FCGR2A* haplotype (2A.3), defined by Q27W, H131H, and the RA-associated SNP rs12746613, which showed the strongest association with RA. Multi-omics analyses demonstrated that 2A.3 is associated with reduced expression of the soluble *FCGR2A* splice variant and lower circulating soluble FcγRIIa levels. Functional studies revealed altered IgG interactions and delayed FcγRIIa signal transduction associated with Q27W, while structural analyses found no evidence for stable ectodomain dimerisation. Together, these findings identify 2A.3 as an important functional contributor to RA susceptibility and provide mechanistic insight into how *FCGR2A* variation may influence immune regulation and disease risk in Europeans.

## INTRODUCTION

FcγRIIa is the widest expressed FcγR, found on monocytes, macrophages, neutrophils and platelets and potentially upregulated on activated endothelial cells ^1–3^. Two main splice variants have been described, a full-length transmembrane receptor and a secreted soluble form (sFcγRIIa) lacking the transmembrane domain ^4^. Polymorphism in *FCGR2A*, the gene encoding FcγRIIa, has been associated with susceptibility to numerous autoimmune pathologies, including rheumatoid arthritis (RA) ^5,6^, systemic lupus erythematosus (SLE) ^7–9^, Takayasu arteritis ^10^, ulcerative colitis (UC) ^11,12^, Kawasaki disease ^13^ and Graves’ disease ^14^. The best characterised *FCGR2A* polymorphism encodes either Arginine or Histidine at amino acid position 131, in the second extracellular receptor domain, in the IgG binding site, hereafter H131R. This polymorphism has long been described as a genetic factor influencing susceptibility and progression/severity of infections including pneumococcal ^15^, meningococcal ^16^, malaria ^17^, dengue ^18^, severe acute respiratory syndrome coronavirus 1 (SARS-CoV-1) ^19^ and 2 ^20^, and respiratory syncytial virus (RSV) ^21^.

The allele conferring susceptibility to different autoimmune disorders and infections seems to be dependent on immune complex composition and cohort ethnicity. In general, GWAS and candidate gene association studies have attributed the observed *FCGR2A* associations to H131R (rs1801274). X-ray structures from FcγRIIa ectodomains co-crystallised with IgG1 have defined the structural basis for the functional effect of H131R, accounting for the high/low responder phenotype observed in human PBMC assays ^22^. The 131H allotype exhibits stronger binding to human than 131R. X-ray crystallographic studies have previously suggested FcγRIIa dimerisation may facilitate signal transduction ^23^, with divergent dimer structures adopted by the 131H and 131R allotypes ^24^. The lower frequency *FCGR2A*-Q27W (hereafter Q27W) polymorphism has attracted less attention but has been described to delay calcium mobilisation on crosslinking of FcγRIIa in patients with hypogammaglobulinemia ^25^. The soluble form of FcγRIIa is reported to be secreted rather than cleaved and plasma concentration is correlated with the Q27W polymorphism ^26^.

Whilst *FCGR2A* is recognised as a susceptibility locus for RA, fine-mapping studies have not been performed to identify the underlying functional variants. In RA, the associated *FCGR2A* variant (rs12746613) ^5^ lies 8.3kb upstream of *FCGR2A* on the telomeric flank of a segmental duplication (SD), which incorporates the majority of the *FCGR* locus. The coverage of high-throughput genotyping technologies is limited in regions of SD. The *FCGR* locus on chromosome 1 is an example of one such region, containing 5 genes in two classes: *FCGR2A*, *FCGR2B*, *FCGR2C*, *FCGR3A* and *FCGR3B*. Copy number variation (CNV) is observed with *FCGR3A*, *FCGR3B* and *FCGR2C*, but not *FCGR2A* or *FCGR2B* (**Figure 1A**) ^27^. Strong sequence identity between the three *FCGR2* genes (*A*, *B* and *C*) has led to erroneous NCBI dbSNP entries for underlying sequence variants. In homologous regions single nucleotide differences (known as paralogous sequence variants, can be used to discriminate one gene from another and to amplify specific genes by PCR, allowing the determination of gene specific polymorphisms to inform the partitioning of variants. Careful gene-specific resequencing could help establish firm anchors for alignments of short sequences generated by parallelised sequencing technologies.

**Figure 1.**
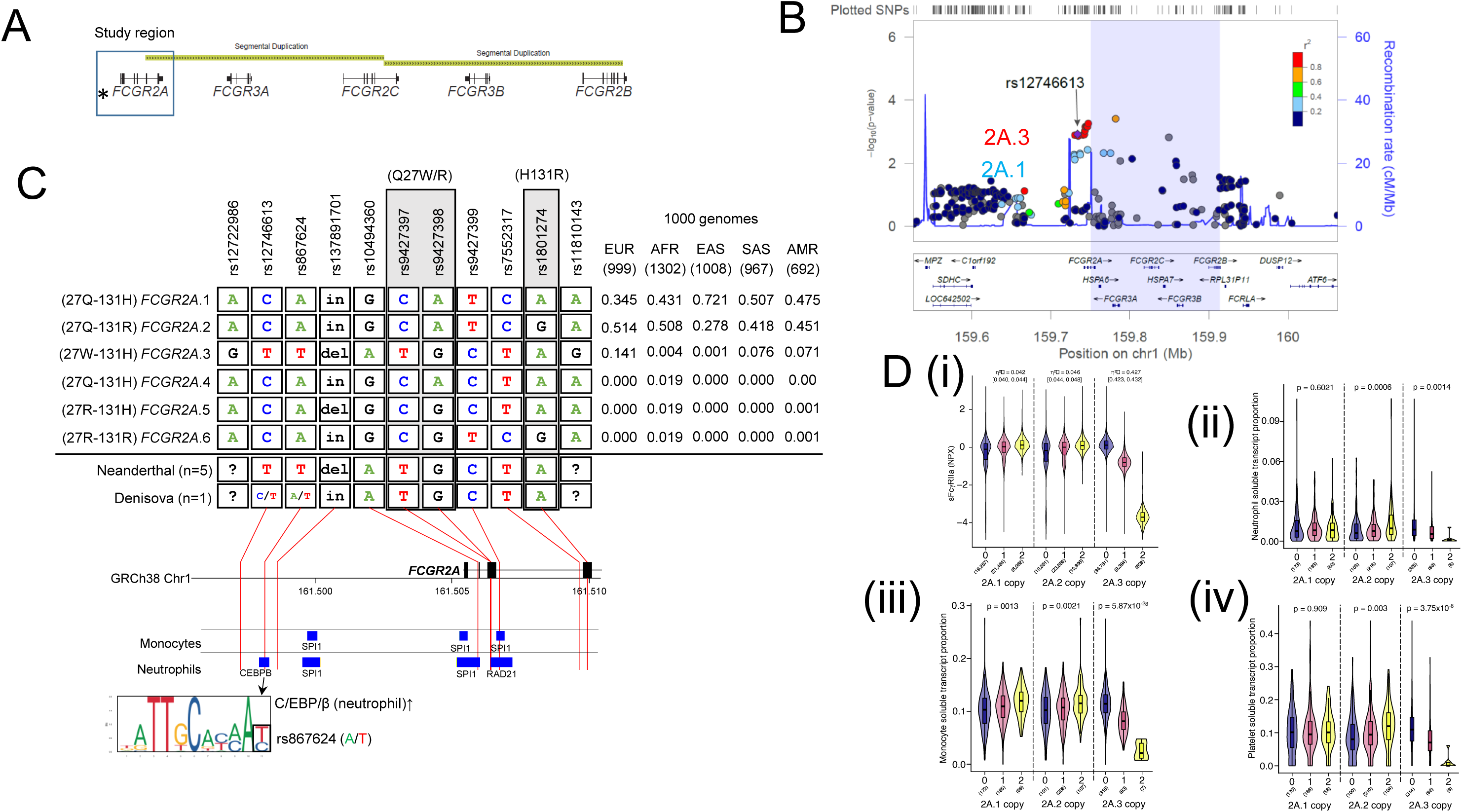
Fine mapping the *FCGR2A* rheumatoid arthritis association. **(A)** The *FCGR* locus on chromosome 1. *Index SNP rs12746613. Segmentally duplicated region indicated in yellow and the study region boxed. **(B)** LocusView plot of single marker associations from Immunochip data in UK RA compared with healthy controls. Segmental duplication indicated by blue shading. **(C)** Functional haplotype frequencies in 1000 genomes European (EUR), African (AFR), East Asian (EAS), South Asian (SAS) and Admixed American (AMR) populations, where numbers in brackets indicate the number of chromosomes analysed. Corresponding nucleotides observed in Archaic hominins (Neanderthal: Vindija x 3; Altai x1; Chagyrskaya x 1; Denisova x 1). 2A.3 haplotype variants mapping to ChIP Atlas transcription factor binding sites active in primary monocytes and neutrophils suggest allele-specific regulation of the 27W-131H allotype in immune cell subsets. **(D)** Soluble FcγRIIa expression. **(Di)** UK Biobank Olink plasma NPX abundance of soluble FcγRIIa grouped by copy number of European haplotypes. Kruskal-Wallis effect sizes (η²ᴴ) with 95% CI and number of individuals for each tested haplotype are in brackets. **(Dii)**NIHR BioResource Rare Disease Phenotyping study splice QTL violin plots of the proportion of *FCGR2A* transcripts lacking exon 5 in neutrophils, platelets **(Diii)** and monocytes **(Div)**, grouped by copy number of each *FCGR2A* haplotype (numbers in each group in brackets), ANOVA p-values calculated using linear regression. LD: linkage disequilibrium, IgG: immunoglobulin G, Fc: fragment crystallisable domain of IgG, QTL: quantitative trait locus, NPX: Normalised protein expression.

We adopted a multi-step approach to fine mapping the *FCGR2A* genetic association in RA: (i) resequencing *FCGR2A*, *FCGR2B* and *FCGR2C* to confirm genuine common variants and calculation of pairwise linkage disequilibrium (LD), followed by *FCGR2A* tag single nucleotide polymorphism (SNP) selection and genotyping of the reduced set of markers on a multiplexed genotyping platform; (ii) re-evaluation of high density SNP typing – Immunochip (custom Illumina Infinium HD array); (iii) combined statistical analysis to identify RA–associated variants in independent data sets; (iv) evaluation of functional haplotypes in *FCGR2A* splicing and protein expression; (v) recombinant reconstruction and production of commonly observed FcγRIIa allotypes as isolated ectodomains, to determine the functional effect of identified nonsynonymous polymorphisms on receptor structure/function relationships; (vi) crystallographic structure determination of the 27W-131H ectodomain; (vii) evaluation of full-length FcγRIIa function using cell-based platforms for measuring IgG subclass preferences and complex interactions; (viii) cellular reporter assays to interrogate the functional impact of nonsynonymous polymorphisms on signal transduction.

## RESULTS

### *FCGR2A* resequencing

We used locus-specific resequencing to measure LD across the SD boundary. Long-specific PCR (*FCGR2A*, *FCGR2B* and *FCGR2C*) and subsequent Sanger sequencing of nested PCR products yielded full sequencing coverage in a panel of 28 Europeans for 21kb of *FCGR2A*, including 8kb of the promoter region, 16.7kb of *FCGR2B* and 20.4kb of *FCGR2C*, enabling discrimination of PSVs and calculations of pairwise LD between all common SNPs including those spanning the *FCGR2A* SD boundary (**Supplementary Figure 1A**). We resolved the underlying gene-specificity of eight SNPs mapping to non-unique regions (**Supplementary Table 1**). Sixty six *FCGR2A* dbSNP records were confirmed in the resequenced region, including two previously reported nonsynonymous coding polymorphisms, Q27W and H131R. The Q27W substitution was encoded by two adjacent but independent dbSNPs (rs9427397 and rs9427398; **Supplementary Figure 1B**). In our European resequencing panel, we only observed the dinucleotide CA>TG substitution.

### Association of *FCGR2A* SNPs with rheumatoid arthritis

We initially used a haplotype tag SNP approach to design a custom Sequenom genotyping array which was used to genotype UK European RA patients (n=1959) and controls (n=1120). Since no direct assay for rs1801274 (H131R) was compatible with the Sequenom panel, we used rs12139150 as a proxy (D’=0.975, r^2^=0.864 in 1000 genomes EUR). We reanalysed additional UK European Immunochip RA/control cohort data (n=3869/8430, respectively) (**Figure 1B**). Eight SNPs were common to both the Sequenom plex and Immunochip (**Supplementary Figure 1C**), with a genotype concordance rate of 97.9-99.8% in 518 overlapping individuals (**Supplementary Figure 1D**). The boundaries of the SD closely corresponded to SNPs that departed from Hardy-Weinberg equilibrium (HWE) among the 544 Immunochip *FCGR* locus SNPs analysed (**Supplementary Figure 2A**). Immunochip SNPs that were obviously affected by CNV in their probe targets were discarded after manually inspecting B-allele frequency (BAF) plots for extra or shoulder peaks, which was included in our quality control (QC) pipeline (**Supplementary Figure 2B**).

**Figure 2.**
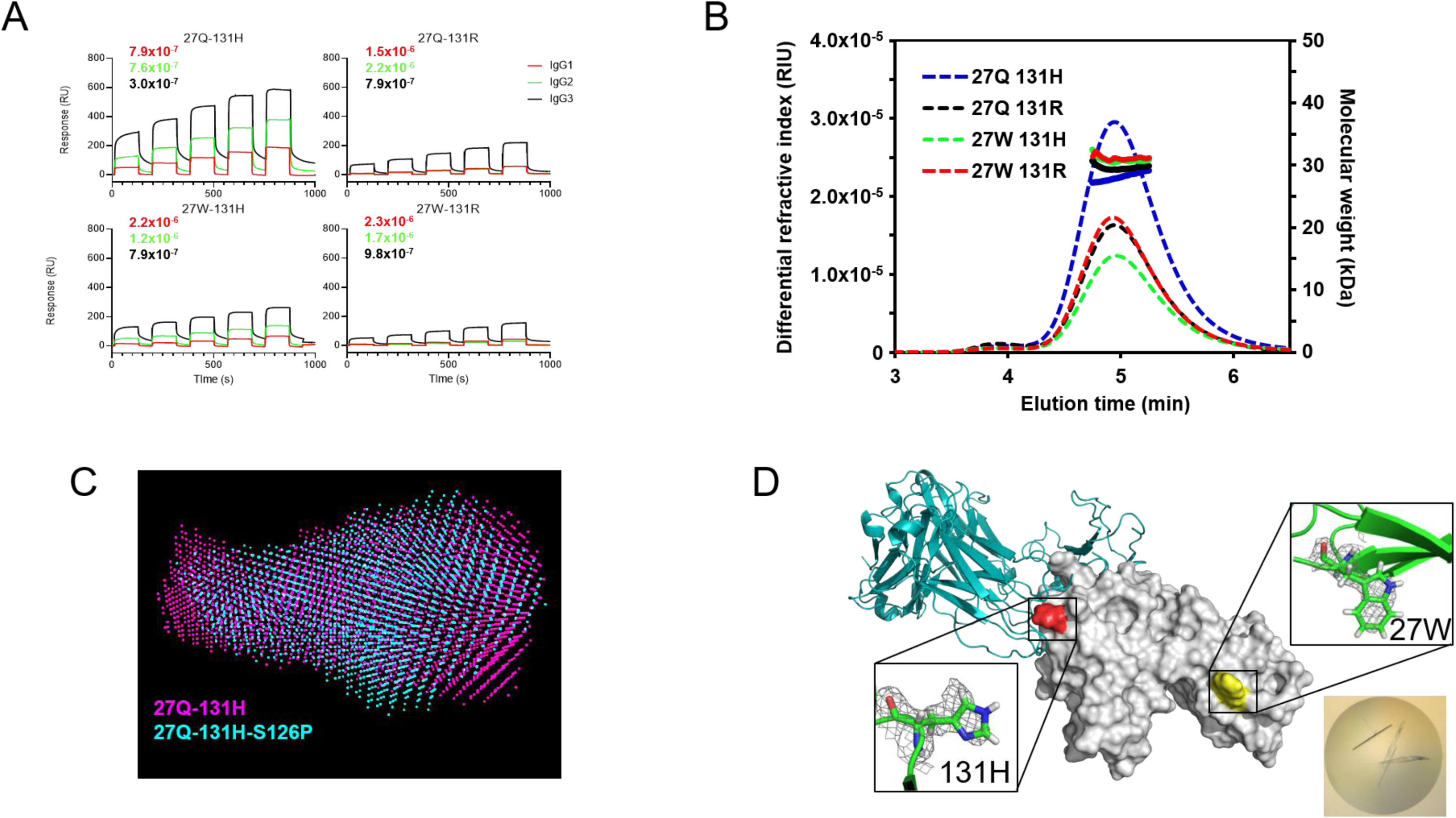
Biophysical and structural impact of *FCGR2A* polymorphisms on soluble FcγRIIa ectodomains. **(A)** Surface plasmon resonance sensorgrams of the interaction between immobilised FcγRIIa ectodomains captured using an anti His murine antibody and soluble monomeric IgG subclasses 1-4 as analytes. Single cycle kinetics of double referenced sensorgrams. On/off rates were too fast to obtain reliable interaction kinetics, molar *K*_D_ values were calculated from steady state for IgG1-3. **(B)** SEC-MALLS elution profiles for four allotypes of soluble FcγRIIa ectodomains conferred by Q27W and H131R. These were the same ectodomains captured on the SPR chips in panel A. **(C)** High-performance liquid chromatography with small angle X-ray scattering (HPLC-SAXS) *ab initio* envelope overlays for 27Q-131H (magenta) and 27Q-131H-S126P (cyan). Both variants eluted at the same volume from the HPLC column. **(D)** X-ray crystallographic model of the 27W-131H allotype (PDB 8CHA) with electron density fitting around the polymorphic residues Q27W (yellow) and H131R (red). Superposed IgG Fc binding position from PDB model 3RY6 represented in teal. Inset crystal image after 5.5 days in 0.1M sodium phosphate citrate, pH4.2 with 41% (v/v) PEG300.

All 13 *FCGR2A* SNPs from the Sequenom Plex and the 25 SNPs on Immunochip that passed BAF QC were included in a combined association analysis of UK European RA (n=5310) and controls (n=9550). Eleven associated markers were in strong LD with rs12746613 (all r^2^>0.97, minor allele frequency (MAF) ~0.12; e.g. rs6671753: *P*=0.0004, adds ratio (OR) [95% confidence interval (CI)] 1.15 [1.06-1.24]; **Table 1**). In UK Europeans, rs12746613 was a perfect proxy for rs9427397, one of the two variants defining Q27W in Europeans. Three variants, with MAF ~0.5, were in LD (r^2^=0.89) (e.g. rs1801274: *P*=0.004, OR [95%CI)] 1.08 [1.03-1.14]). When a marker tagging each functional variant (Q27W and H131R) was included in the same multiple regression model, the effect of both markers was attenuated, but showed some evidence of independent association (Q27W (rs9427399) *P*=0.022, OR [95%CI] 1.11 [1.01-1.21] and H131R (rs1801274) *P*=0.087, OR [95%CI] 1.05 [0.99-1.12]).

**Table 1.**
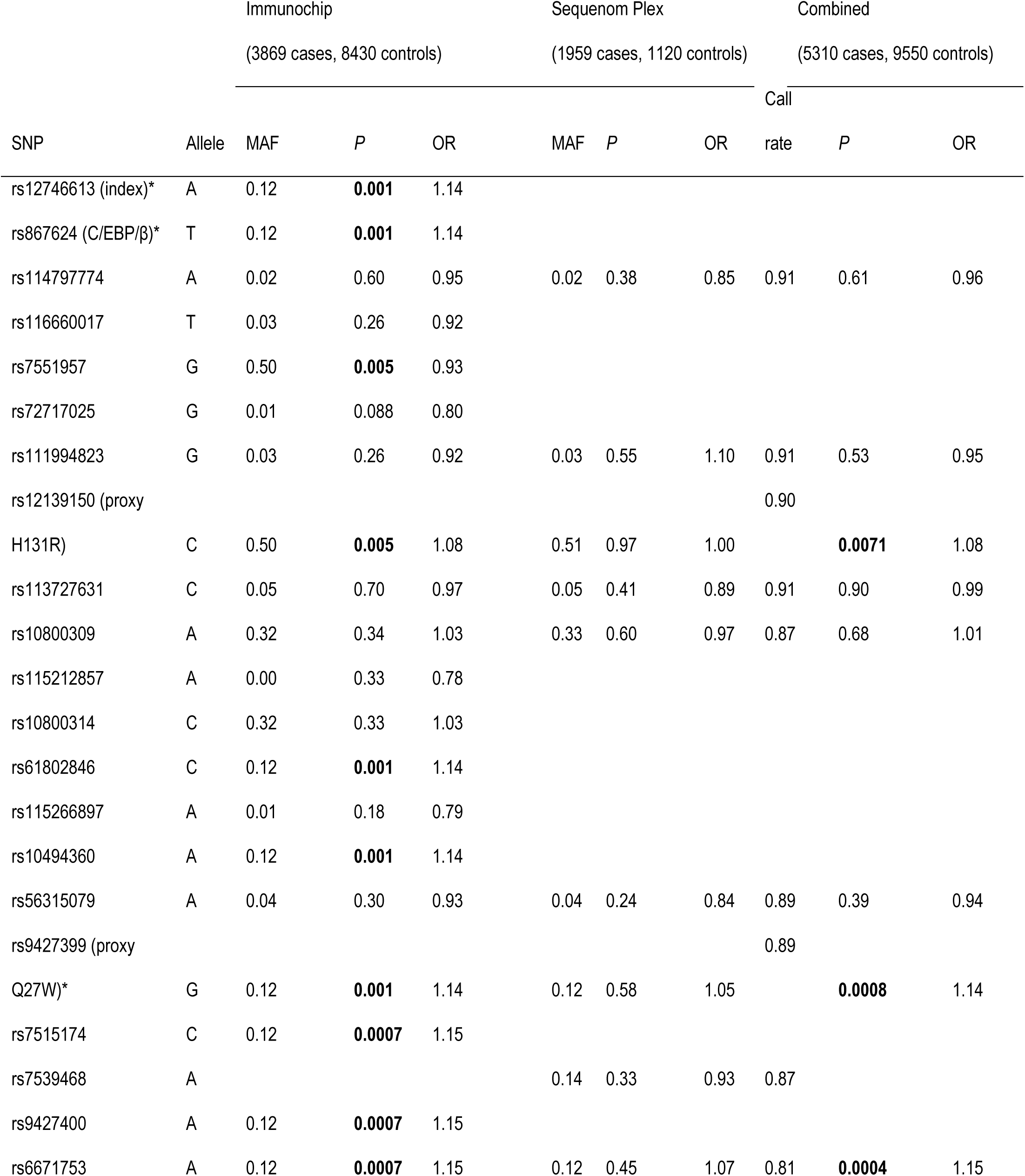
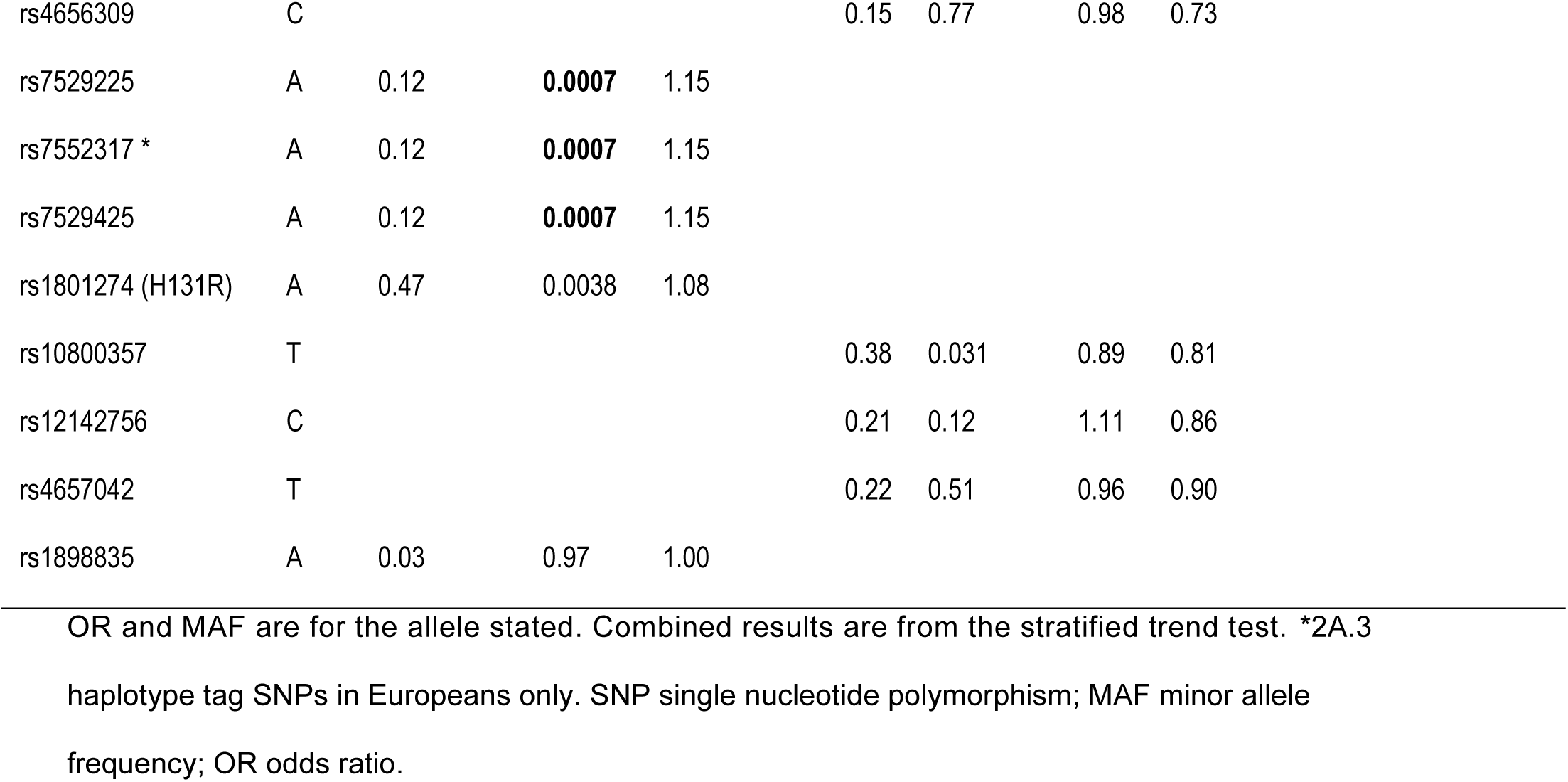
Trend test results for the association of rheumatoid arthritis with SNPs in the *FCGR2A* region for the Immunochip and Sequenom plex data.

| SNP | Allele | Immunochip<br>(3869 cases, 8430 controls) |  |  | Sequenom Plex<br>(1959 cases, 1120 controls) |  |  | Combined<br>(5310 cases, 9550 controls) |  |  |
| --- | --- | --- | --- | --- | --- | --- | --- | --- | --- | --- |
|  |  | MAF | *P* | OR | MAF | *P* | OR | Call<br>rate | *P* | OR |
| rs12746613 (index)* | A | 0.12 | **0.001** | 1.14 |  |  |  |  |  |  |
| rs867624 (C/EBP/β)* | T | 0.12 | **0.001** | 1.14 |  |  |  |  |  |  |
| rs114797774 | A | 0.02 | 0.60 | 0.95 | 0.02 | 0.38 | 0.85 | 0.91 | 0.61 | 0.96 |
| rs116660017 | T | 0.03 | 0.26 | 0.92 |  |  |  |  |  |  |
| rs7551957 | G | 0.50 | **0.005** | 0.93 |  |  |  |  |  |  |
| rs72717025 | G | 0.01 | 0.088 | 0.80 |  |  |  |  |  |  |
| rs111994823 | G | 0.03 | 0.26 | 0.92 | 0.03 | 0.55 | 1.10 | 0.91 | 0.53 | 0.95 |
| rs12139150 (proxy<br>H131R) | C | 0.50 | **0.005** | 1.08 | 0.51 | 0.97 | 1.00 | 0.90 | **0.0071** | 1.08 |
| rs113727631 | C | 0.05 | 0.70 | 0.97 | 0.05 | 0.41 | 0.89 | 0.91 | 0.90 | 0.99 |
| rs10800309 | A | 0.32 | 0.34 | 1.03 | 0.33 | 0.60 | 0.97 | 0.87 | 0.68 | 1.01 |
| rs115212857 | A | 0.00 | 0.33 | 0.78 |  |  |  |  |  |  |
| rs10800314 | C | 0.32 | 0.33 | 1.03 |  |  |  |  |  |  |
| rs61802846 | C | 0.12 | **0.001** | 1.14 |  |  |  |  |  |  |
| rs115266897 | A | 0.01 | 0.18 | 0.79 |  |  |  |  |  |  |
| rs10494360 | A | 0.12 | **0.001** | 1.14 |  |  |  |  |  |  |
| rs56315079 | A | 0.04 | 0.30 | 0.93 | 0.04 | 0.24 | 0.84 | 0.89 | 0.39 | 0.94 |
| rs9427399 (proxy<br>Q27W)* | G | 0.12 | **0.001** | 1.14 | 0.12 | 0.58 | 1.05 | 0.89 | **0.0008** | 1.14 |
| rs7515174 | C | 0.12 | **0.0007** | 1.15 |  |  |  |  |  |  |
| rs7539468 | A |  |  |  | 0.14 | 0.33 | 0.93 | 0.87 |  |  |
| rs9427400 | A | 0.12 | **0.0007** | 1.15 |  |  |  |  |  |  |
| rs6671753 | A | 0.12 | **0.0007** | 1.15 | 0.12 | 0.45 | 1.07 | 0.81 | **0.0004** | 1.15 |

|  |  |  |  |  |  |  |  |  |
| --- | --- | --- | --- | --- | --- | --- | --- | --- |
| rs4656309 | C |  |  |  | 0.15 | 0.77 | 0.98 | 0.73 |
| rs7529225 | A | 0.12 | **0.0007** | 1.15 |  |  |  |  |
| rs7552317 * | A | 0.12 | **0.0007** | 1.15 |  |  |  |  |
| rs7529425 | A | 0.12 | **0.0007** | 1.15 |  |  |  |  |
| rs1801274 (H131R) | A | 0.47 | 0.0038 | 1.08 |  |  |  |  |
| rs10800357 | T |  |  |  | 0.38 | 0.031 | 0.89 | 0.81 |
| rs12142756 | C |  |  |  | 0.21 | 0.12 | 1.11 | 0.86 |
| rs4657042 | T |  |  |  | 0.22 | 0.51 | 0.96 | 0.90 |
| rs1898835 | A | 0.03 | 0.97 | 1.00 |  |  |  |  |

OR and MAF are for the allele stated. Combined results are from the stratified trend test. \*2A.3
haplotype tag SNPs in Europeans only. SNP single nucleotide polymorphism; MAF minor allele frequency; OR odds ratio.

Previous associations at the *FCGR* locus have been more pronounced in autoantibody positive subgroup analyses ^28,29^. However, neither Q27W nor H131R conferred notable differences in ORs (Q27W: rs9427399 anti-citrullinated peptide antibody (ACPA)+ve *P*=0.001, OR [95% CI] 1.17 [1.06-1.28]; ACPA-ve *P*=0.083, OR [95% CI] 1.13 [0.98-1.29] and H131R: rs1801274 ACPA+ve *P*=0.029, OR [95% CI] 1.07 [1.00-1.14]; ACPA-ve *P*=0.093 OR [95% CI] 1.08 [0.99-1.19]).

### Constructing functional *FCGR2A* haplotypes

To rationalise LD in *FCGR2A,* we constructed haplotypes based on our resequencing-confirmed SNPs, previous reports of their functional qualitative (amino acid substitution) and quantitative (regulation of expression level) effects (JASPAR,^30^), and their selectability based on a selective sweep survey ^31^. We then queried the 1000 genomes (1KG) phase 3 data set to determine worldwide population frequencies of these SNP haplotypes. Phased haplotypes from 999 European chromosomes revealed the dinucleotide *FCGR2A*-Q27W alleles conferred by rs9427397 and rs9427398. In the 1KG African group, a third allele conferred by rs9427398 in isolation, encoding arginine (*FCGR2A*-27R) was present. We subsequently resequenced across exon 3 in 43 individuals of African ancestry and confirmed *FCGR2A*-27R in three individuals (example shown in **Supplementary Figure 1B**). We classified the extended haplotypes based on global frequency, noting low haplotype diversity in East Asians (EAS), where only 2A.1 and 2A.2 were present; three common *FCGR2A* haplotypes (2A.1, 2A.2 and 2A.3) in Europeans (EUR) and South Asians (SAS), and a total of six haplotypes over 1% frequency in Africans (AFR). Of 5008 phased chromosomes, 4968 (99.2%) were assignable to one of the six previously defined *FCGR2A* haplotypes, whereas 40 chromosomes (0.8%) carried non-canonical haplotypes. Of these noncanonical haplotypes 20 were African, 11 South Asian, 7 European and 2 Admixed American. Sequence data for *FCGR2A* from five Neanderthals^32–34^ and one Denisovan^35^ enabled comparison with the modern human haplotypes, albeit with very low coverage. Formal analysis of archaic introgression in *FCGR2A* was downloaded from Arcseqhub^36^ and is summarised in **Supplementary Figure 3**.

**Figure 3.**
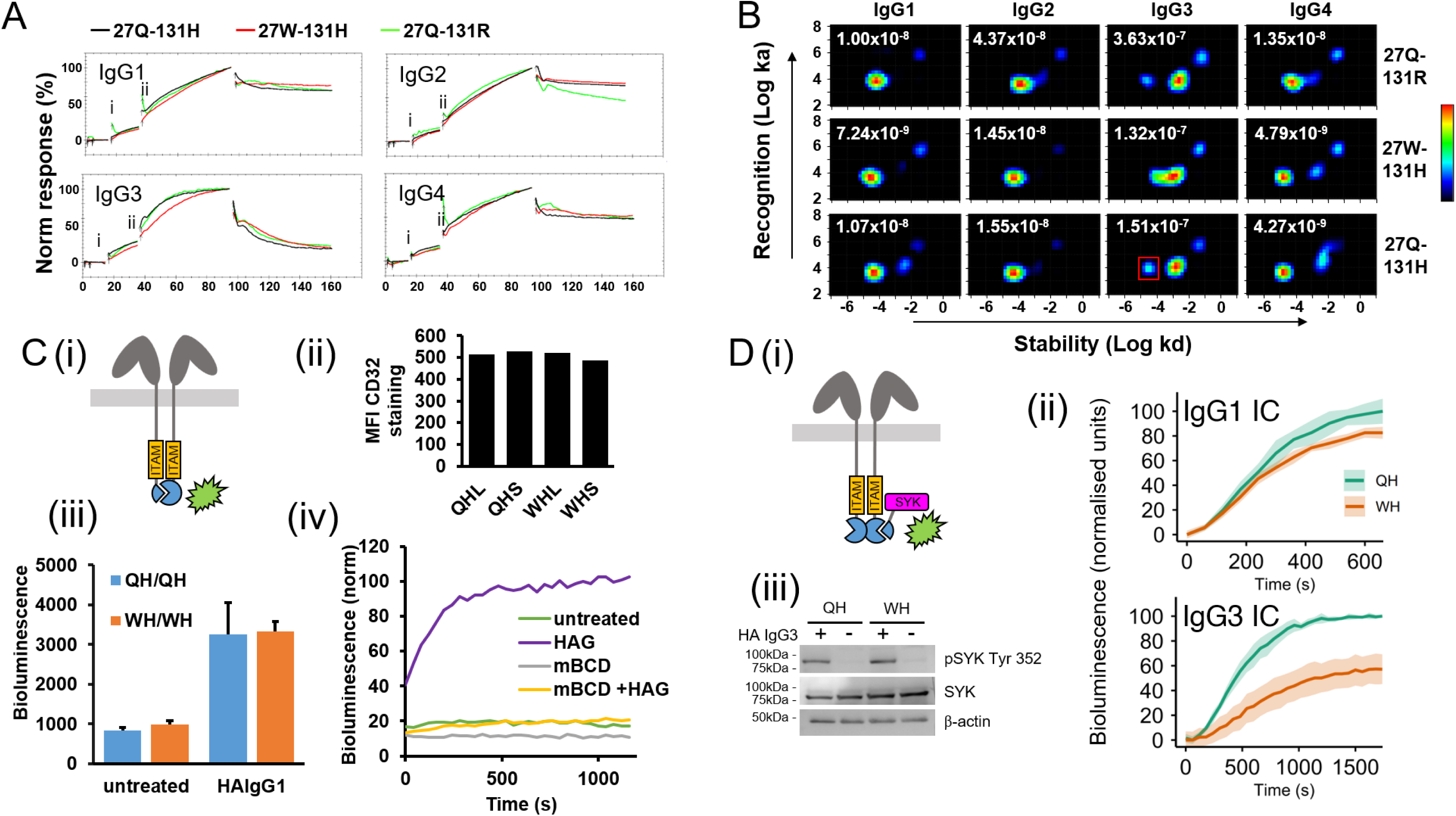
Cellular FcγRIIa – IgG interactions. **(A)** Real time interactions between live cellular-expressed FcγRIIa allotypes and soluble monomeric IgG subclasses 1-4 using Ligand Tracer. Full-length FcγRIIa allotypes were ectopically expressed on Spodoptera frugiperda (Sf9) cells after baculoviral transduction. Labelled (DyLight650) soluble monomeric IgG ligands were added in two stages at 15nM **(Ai)** and 45nM **(Aii)**, before being replaced with PBS at 100 minutes to measure the dissociation phase. Representative examples of triplicate sensorgrams are shown. **(B)** Global Interaction Map plots for each receptor-ligand pair measured by Ligand Tracer. Global interaction calculations were performed on combined triplicate sensorgrams. In each map the dominant interaction molar KD is given. The boxed interaction in the 27Q-131H/IgG3 panel is referenced in the text. **(Ci)** Nanobit luciferase complementation reporter assay for FcγRIIa dimerisation in 27Q-131H and 27W-131H allotypes, where FcγRIIa-SmBiT and FcγRIIa-LgBiT were transiently cotransfected into HEK293 cells. **(Cii)** Expression levels of each reporter assay component were measured by flow cytometry, staining for CD32 with FITC-labelled antibody clones IV.3 and AT-10 in the same tube. L Large bit, S Small bit. **(Ciii)** Bioluminescence in untreated cells represented background luciferase complementation, which rose three-fold upon addition of 50 µg/mL heat aggregated IgG1. Error bars indicate variance over three biological replicates (separate transient transfections). **(Civ)** Addition of 10mM mβCD prevented the formation FcγRIIa-rich signalling domains, with and without HAG1. **(Di)** Time-resolved nanobit assay reporting association of activatory adaptor SYK with the ITAM of FcγRIIa. **(Dii)** Syk association with the ITAM of 27Q-131H and 27W-131H FcγRIIa in response to 75 µg/mL HAG1 and 30 µg/mL HAG3. Bioluminescent signal was generated by QH- and WH-LgBiT - tagged receptors binding SmBiT – SYK, induced by HAG1. Error bands represent standard deviation from biological triplicates. Results shown are representative of five independent experiments. **(Diii)** Western blots demonstrate that HAG3-induced binding of SYK to FcγRIIa ITAM correlated with SYK phosphorylation (Tyr 352). SYK Spleen tyrosine kinase, ITAM Immunoreceptor tyrosine-based activation motif, HAG Heat aggregated IgG.

### Association of *FCGR2A* haplotypes with rheumatoid arthritis

LD between Q27W (proxy rs9427399) and H131R (rs1801274) was strong, (D’=1), since the 27W allele was not observed with 131R in the entire UK data set. However, because of the large difference in frequency between these variants, r^2^ was low at 0.16. In the haplotype trend test using the more prevalent 27Q-131R (2A.2) haplotype as a reference, the haplotype encoding 27W-131H (2A.3) showed the strongest association with RA susceptibility (P=0.0003, OR [95%CI] 1.17 [1.07-1.27]; Supplementary Table 4), whereas 27Q-131H (2A.1) haplotype showed marginal increased RA risk (P=0.087, OR [95% CI] 1.05 [0.99-1.12]). When stratifying for autoantibody status the 27W-131H (2A.3) haplotype conferred a marginally increased risk in ACPA +ve RA OR [95% CI] 1.18 [1.07-1.30], P=0.0007 compared with ACPA −ve RA 1.15 [1.00-1.33], P=0.05.

### *FCGR2A* association is independent of previously reported *FCGR3A*-F158V and *FCGR3B* copy number variation association in rheumatoid arthritis

We previously reported that the *FCGR3A*-F158V (rs396991) variant and reduced *FCGR3B* copy number (CN) were associated with RA ^28,29^. To determine whether the signal in *FCGR2A* could be explained by LD, the markers rs9427399 (Q27W proxy r^2^=0.99) and rs12139150 (H131R proxy r^2^=0.86) genotyped using the Sequenom plex were included in the same models as previously genotyped *FCGR3A*-F158V and *FCGR3B* deletions (n=732 cases and 366 controls). The ORs for the markers remained almost unchanged after adjustment for *FCGR3B* deletion, with minor change after *FCGR3A*-158V, suggesting the associations were independent (**Supplementary Table 2**). LD was evident between *FCGR3B* deletions and the non-risk allele for rs9427399 (D’=0.63), and the non-risk allele for rs1801274 (D’=0.23), further suggesting the CNV does not account for the associations found with either of the two groups of *FCGR2A* SNPs (**Supplementary Table 3**).

### Replication in a second rheumatoid arthritis cohort

Having identified a risk haplotype of 27W-131H (2A.3) in UK European RA we then sought replication in a Spanish European RA cohort of 843 cases and 1953 Spanish controls where Immunochip data were available. As with the UK cohort the 27W-131R haplotype was absent. The 27W-131H haplotype was again associated with RA, with a comparable effect size although with lower statistical significance, most likely a reflection of the smaller sample size (**Supplementary Table 4**).

### Investigation of putative FcγRIIa expression variants

Bioinformatic analyses (chromatin immunoprecipitation - ChIP Atlas ^37^) suggested FcγRIIa expression may be regulated by transcription factors specific for different leukocyte lineages (**Figure 1C**). We searched for signals of a potential expression quantitative trait locus (eQTL) in the Genotype-Tissue Expression Project (GTEx) V8^38^ and found evidence that the 2A.3 haplotype was associated with decreased transcription of a soluble *FCGR2A* splice variant (s*FCGR2A*) in whole blood and tissues (**Supplementary Figure 4**). In tissues expression of *FCGR2A* was confined largely to dendritic cells and macrophages ^39^. To determine whether the 2A.3 haplotype-encoded reduced transcription of s*FCGR2A* leads to detectably lower sFcγRIIa in blood, we interrogated the UK Biobank proteomics data set. In UK Biobank only one 2A.3 haplotype tag SNP (rs12722986) was reported. After deriving copy number of each *FCGR2A* haplotype, we correlated Olink NPX values for 46,803 individuals with corresponding protein expression data. Reflective of the splice QTL observed in GTEx, increasing copy number of the 2A.3 haplotype was strongly associated with lower plasma sFcγRIIa (**Figure 1D(i)**), suggesting that one quantitative functional effect of the RA-associated haplotype is to reduce the plasma concentration of sFcγRIIa. Systematic analysis of the 2A.1 and 2A.2 haplotypes on sFcγRIIa abundance revealed negligible effects.

To investigate the breadth of this 2A.3 spliceQTL/pQTL effect, we expanded our analysis to different cell types by utilising a pre-release snapshot of the NIHR BioResource rare disease RNA phenotyping project data, where RNAseq data were available for separated neutrophils, monocytes, CD4+ T-cells and platelets of 440 individuals, including rare diseases and healthy controls, without selecting ancestry. Here, we analysed bulk RNAseq data by counting the number of s*FCGR2A* transcripts expressed as a proportion of the total number of *FCGR2A* transcripts. As expected, *FCGR2A* transcription in CD4+ T-cells was absent. We found that ~10% of *FCGR2A* transcripts in monocytes encoded the soluble form of FcγRIIa, but in individuals carrying one copy of the 2A.3 haplotype this dropped to ~7%, and ~2% in homozygotes (**Figure 1Diii**). Platelets displayed more profound s*FCGR2A* transcript reduction, starting at 10% and dropping to ~0 in 2A.3 homozygotes (**Figure 1Div**). Similar soluble transcript proportion reductions were observed in neutrophils, but from a lower maximum level of ~1% in the absence of 2A.3 (**Figure 1Dii**). These findings provide data which can be used to evaluate the likely sources of plasma sFcγRIIa in non-2A.3 carriers.

### Effect of nonsynonymous FcγRIIa polymorphisms on IgG subclass binding preferences using soluble ectodomains

We used surface plasmon resonance (SPR) to measure the effect of Q27W and H131R on IgG subclass binding kinetics. We captured soluble His-tagged FcγR ectodomains bearing all four allotypic combinations (27Q-131H, 27Q-131R, 27W-131H and 27W-131R) on a carboxymethylated dextran with an amine coupled murine anti-His tag antibody and then injected serial dilutions of purified IgG subclass solutions from 188 nM to 3 µM as analytes in single cycle kinetics. Kinetic constants were too rapid to be determined due to the lack of sensorgram curvature (**Figure 2A**) and affinity constants were determined through analysis of steady states (**Supplementary Table 5**). We confirmed the selective preference of the 131H ectodomain for IgG2 over 131R and increased binding affinity for IgG3 compared to 131R, irrespective of the Q27W variant. Interactions with IgG4 subclass (not shown) did not reach steady state, indicating possible complex kinetics, aggregation or nonspecific binding.

### Effect of FcγRIIa polymorphisms on recombinant soluble ectodomain dimerisation

In view of reported FcγRIIa ectodomain dimerisation ^24^, and to assess the potential for Q27W in modulating such interactions, we sought to confirm this observation using multiple biophysical methods. We subjected the purified FcγRIIa ectodomains to size exclusion chromatography with multi-angle laser light scattering (SEC-MALLS) to determine their quaternary state. All ectodomain allotypes appeared to exist as monomers at the concentrations analysed (**Figure 2B**). We then attempted to detect FcγRIIa dimers by comparing the wild-type FcγRIIa (27Q-131H) ectodomain with a version carrying the S126P mutation previously reported to prevent receptor dimerisation ^40^. We synthesised 27Q-131H and 27Q-131H-S126P in HEK293T cells by transient transfection and subjected these to high-performance liquid chromatography with small angle X-ray scattering (SEC-SAXS). Both variants eluted at the same time and X-ray scattering data collected prior to accumulation of radiation damage enabled the calculation of *ab initio* envelopes which could be overlaid, thus demonstrating there was no difference in oligomeric state (**Figure 2C**).

We then sought to determine the effect of Q27W on the FcγRIIa ectodomain fold and the formation of crystallographic (artefactual) or biologically relevant oligomerisation. We crystallised and resolved the structure of the 27W-131H allotype (**Figure 2D**, PDB 8CHA, **Supplementary Table 6**) and compared the model with PDB 3RY4 (27Q-131H) ^24^. There was no evidence of fold disruption or dimerisation, crystallographic or otherwise, in the 27W-131H allotype. Systematic ClusPro-DC analysis ^41^ of a re-refined 3RY4 model suggested that any dimers were highly likely to be crystallographic artefacts rather than biologically relevant. Unable to reproduce plausible oligomeric states in isolated FcγRIIa ectodomains, we reasoned that any dimerisation activity was likely to be transient and dependent on transmembrane and/or intracellular contacts, so any functional effect of the Q27W polymorphism would only be seen using full-length FcγRIIa in a membrane.

### Real-time measurement of FcγRIIa interactions with IgG subclasses in live cells

To test the assertion that immobilised FcγRIIa ectodomains in SPR may not interact with ligands in the same manner as full-length receptors in a cellular membrane, we expressed the three observed allotypes of FcγRIIa in *Spodoptera frugiperda* (Sf9) cells by baculoviral transduction. We then captured these cells in a Ligand Tracer (LT) Green instrument and monitored their interaction kinetics with fluorescently labelled, uncomplexed IgG subclasses. Interaction kinetics were notably slower in the cellular system when compared to the same interacting pair in SPR. However, the IgG subclass preferences of FcγRIIa allotypes were consistent across both platforms, where the 131H allotype recognised IgG2 more strongly than 131R (**Figure 3A**, **Supplementary Table 5**). Additionally, LT analysis demonstrated a slower association rate for IgG3 from 27W-131H compared to other receptor allotypes. It was also noteworthy that in SPR IgG4 failed to reach steady state on all FcγRIIa allotypes, indicating departure from a 1:1 Langmuir model. In the LT analysis, IgG3 and IgG4 – FcγRIIa interactions conformed to a 1:2 model, suggesting interaction heterogeneity. We applied a global interaction analysis (Interaction Map, IM) to the LT sensorgrams in view of the potential complexity of cell-expressed FcγRIIa interactions and the multiple conformations postulated in the literature. This analysis method is not constrained by the assumption of a 1:1 or 1:2 model but generates an interaction map of individual binding events that contribute to the observed sensorgram (**Figure 3B**). In contrast to SPR, the LT IMs highlighted a difference in potential higher order interactions between receptors and IgG3, where a higher affinity sub-interaction component seen with 27Q was abolished in the 27W interaction map, possibly merged with the main interaction. This analysis also highlighted main interaction affinity enhancement conferred by 27W-131H for IgG1 (~1.5 fold) compared to 27Q-131H, and by 131H for IgG4 (~3 fold) compared to 131R. It is not possible to infer the biological mechanisms from peaks on LT interaction maps and we therefore investigated the effect of this polymorphism on quaternary state and signal transduction in cells.

### Full-length FcγRIIa dimerisation and signal transduction

We investigated the effect of Q27W on oligomerisation and intracellular signal transduction using NanoBiT complementation assays (Promega, Madison, WI). We constructed vectors based on pFC34K LgBiT TK neo that expressed full-length FcγRIIa-27Q and 27W on the FcγRIIa-131H background tagged with a LgBiT on the C terminal, and a vector based on pFC36K SmBiT TK neo, expressing FcγRIIa-27Q and -27W with SmBiT tagged on the C terminal immunoreceptor tyrosine-based activation motif (ITAM) (**Figure 3Ci**). Transfection of both Q27W allotypic vectors into HEK293 cells allowed spontaneous and immune complex-induced close association/dimerisation of the receptors to be studied. We used flow cytometry to ensure that both Q27W allotypes were expressed at similar levels in all transfections (**Figure 3Cii**) and further validated by measuring the peak height of bioluminescence in transient and stably transfected cells (see below). Bioluminescence due to complementation of the nanoluc enzyme was detectable at low levels in resting cells, rising three-fold in cells treated with heat-aggregated IgG1 (HAG1) as a surrogate for immune complexes. Q27W had no effect on the level of resting or HAG1-induced bioluminescence (**Figure 3Ciii**). On addition of the cholesterol-chelating drug methyl-β-cyclodextrin (MβCD), the bioluminescence due to HAG1 crosslinking was suppressed, as was the resting signal, indicating the importance of cell membrane signalling microdomains for transient FcγRIIa oligomerisation (**Figure 3Civ**).

To investigate the potential impact of Q27W on signal transduction, we created a nanobit assay containing SmBiT tethered to the N-terminus of spleen tyrosine kinase (SYK), to report the interaction between SYK and the ITAM of FcγRIIa. Coupled with the existing FcγRIIa-LgBiT assay components we were able to measure the association of SYK in real time with the ITAM of 27Q and 27W in response to IgG cross linking (**Figure 3Di**). Cell lines expressing equivalent amounts of the FcγRIIa 27Q and 27W /SYK reporter assays (**Figure 3Dii**) were simultaneously stimulated with defined IgG1-containing immune complexes (upper panel) and the bioluminescent response due to SYK association with the FcγRIIa ITAM was monitored over time. Background unstimulated signals were comparable between 27Q and 27W (n=15 each, P=0.11 Rank Sum Test); however, in response to IgG1 immune complexes (anti FITC clone 4M5.3 hIgG1 complexed with BSA-FITC) the ITAM:SYK association increased more rapidly for 27Q and achieved approximately 20% more signal intensity of the 27W allotype after 10 minutes. Stimulation with immune complexes containing IgG3 (anti ox-LDL clone E06 hIgG3 complexed with oxidised LDL) resulted in delayed signalling of half the intensity for 27W expressing cells compared with 27Q (**Figure 3Dii**). In the same reporter cell lines, on stimulation with 50 µg/mL HAG3, both allotypes transduced intracellular signals resulting in SYK phosphorylation at Tyrosine 352, as demonstrated by Western blotting, where similar levels of phosphorylation were observed for each allele after 300 seconds of stimulation (**Figure 3Diii**).

## DISCUSSION

### Fine mapping of the rheumatoid arthritis association at *FCGR2A*

Historically, disease associations at *FCGR2A* have largely been attributed to the H131R polymorphism because of its well-established effects on IgG subclass binding and its strong linkage disequilibrium with surrounding variants. However, extensive LD at the *FCGR* locus^42^, together with the challenges of analysing a region embedded within segmental duplication, has hindered identification of the variants directly responsible for disease susceptibility. By combining locus-specific resequencing with large-scale genetic association analyses, we demonstrate that the strongest RA association is conferred by the 2A.3 haplotype, which includes Q27W together with multiple variants mapping to regulatory elements. These findings suggest that *FCGR2A*-associated disease risk is unlikely to be explained by a single polymorphism and instead reflects the combined effects of coding and regulatory variation acting within an extended functional haplotype.

Replication of the association in an independent Spanish cohort supports the robustness of this observation and suggests that 2A.3 contributes to RA susceptibility across European populations. While previous studies have implicated neighbouring *FCGR3A*^29^ and *FCGR3B*^28^ variants in autoimmune disease susceptibility, our analyses indicate that the *FCGR2A* association cannot be explained solely by linkage disequilibrium with *FCGR3B* copy number variation. The modest relationship observed between 2A.3 and *FCGR3A*-158V further suggests that multiple susceptibility alleles within the broader *FCGR* locus may contribute independently to disease risk.

Together, these findings support the view that future studies of the *FCGR* locus should focus on functional haplotypes rather than individual variants. The scarcity of 2A.3 outside European populations also suggests that disease associations attributed to *FCGR2A* in different ancestral groups may not share the same underlying molecular basis, emphasising the importance of population-specific fine mapping.

### Population diversity and evolutionary history of *FCGR2A* haplotypes

Our haplotype analyses revealed striking population differences in *FCGR2A* diversity. Europeans and South Asians carried three common functional haplotypes, East Asians only two, whereas Africans harboured six common haplotypes, consistent with the greater genetic diversity and deeper evolutionary history of African populations^43,44^, with some evidence of 2A.5 and 2A.6 in Admixed Americans. Notably, we identified an African-specific *FCGR2A*-27R allele that, despite being present in the 1KG dataset, has not previously been recognised. This appears to result from the independent segregation of rs9427397 and rs9427398, the two SNPs comprising the Q27W dinucleotide polymorphism, which occurs independently of H131R in African populations and expands the repertoire of functional haplotypes observed elsewhere. Conversely, the complete absence of the 27W-131R across all populations analysed suggests that 27W is trapped within the 2A.3 haplotypic background. Together, these observations indicate that *FCGR2A* has undergone a more complex evolutionary history than is apparent from studies restricted to European populations.

Comparison with archaic hominin genomes, together with formal introgression analyses based on the T2T-CHM13 reference genome^36^, suggests that 2A.3 forms part of an Archaic derived haplotype present predominantly in Europeans and to a lesser extent South Asian populations. This interpretation is consistent with growing evidence that introgressed immune loci have contributed to variation in host defence and disease susceptibility in modern humans through effects on both innate and adaptive immunity^45,46^. The scarcity of 2A.3 in East Asian populations may contribute to the lack of reproducible *FCGR2A* associations reported for this ancestry.

The convergence of autoimmune association, immune-regulatory variation and archaic ancestry places *FCGR2A* among a growing number of introgressed immune loci with demonstrable effects on contemporary human phenotypes. Recent work has shown that Neanderthal-derived haplotypes at the immunoglobulin heavy-chain (IGH) locus influence IgG biology and autoimmune disease susceptibility, raising the possibility that multiple components of the antibody-Fcγ receptor axis have been shaped by archaic introgression and subsequent pathogen-driven selection ^47^. Consistent with this interpretation, the persistence of the 2A.3 haplotype despite its association with autoimmune disease may reflect antagonistic pleiotropy, whereby variants that enhance protection against infection are maintained despite increasing susceptibility to inflammatory disease ^48^. In this context, the observed effects of Q27W on FcγRIIa interactions involving IgG3 are of particular interest, as IgG3 is a key mediator of antiviral immunity. Historical advantages in host defence may therefore have contributed to the retention of this introgressed haplotype within European populations despite its detrimental effects on autoimmune disease risk.

#### Quantitative effects on FCGR2A expression

Our multi-omics analyses indicate that one of the most important functional consequences of the 2A.3 haplotype is altered regulation of *FCGR2A* splicing. The consistent direction of effect observed across genomic, transcriptomic and proteomic datasets strongly supports a genuine biological mechanism rather than a platform-specific artefact. The reduction in circulating sFcγRIIa associated with the RA-risk haplotype may have important immunological consequences. Soluble FcγRIIa inhibits the Arthus reaction^49^ and has previously been proposed to act as a decoy receptor capable of modulating immune-complex driven inflammation^50^. Reduced endogenous production of sFcγRIIa could therefore increase local availability of pathogenic immune complexes, favouring activation of cell-surface Fcγ receptors. Such a mechanism is attractive in RA, where immune-complex signalling is believed to contribute to synovial inflammation and tissue damage^51^.

The functional consequences of 2A.3 are not restricted to receptor coding variation and splicing but extend to regulation of *FCGR2A*. Consistent with this interpretation, rs10494360, a variant carried on the 2A.3 haplotype, has previously been identified as part of a functional regulatory element controlling FcγRIIa expression^52^, providing a potential third aspect of functional significance.

Our findings highlight the need to distinguish between total *FCGR2A* expression and isoform-specific regulation. Regulatory variants within the extended 2A.3 haplotype may influence receptor biology not simply through changes in overall transcription but through altering the balance between membrane-bound and soluble receptor forms. Future studies should address how these variants affect receptor abundance in individual immune-cell subsets during inflammation.

These observations may also have translational implications. Both haplotype copy number and circulating soluble FcγRIIa concentration warrant investigation as biomarkers for therapies targeting FcγR pathways. Such measures could potentially prove useful in predicting therapeutic response or informing dose selection for FcγR-directed biologics.

#### Functional effects of Q27W

To assess the functional significance of the *FCGR2A* Q27W and H131R polymorphisms, we generated all four variant combinations as both soluble ectodomains and full-length cellular receptors, enabling complementary biophysical and cellular analyses of IgG binding and signal transduction.

### Isolated ectodomains

SPR analysis confirmed previously reported IgG subclass preferences associated with FcγRIIa-131H and -131R^53^. Although absolute affinities were stronger than those reported previously, likely reflecting methodological differences, our approach enabled direct comparison of receptor binding capacity. The principal finding was that the Q27W polymorphism had little effect on the established H131-associated preference for IgG2, whereas the 27Q-131H allotype showed substantially greater IgG3 binding capacity than 27Q-131R. Using this reductionist approach, we were able to examine receptor-ligand interactions in isolation, however the absence of a membrane environment limits its capacity to capture receptor organisation and signalling processes that may depend on cellular context.

Our second objective was to examine reports that FcγRIIa ectodomains form functional dimers^24^. Using SEC-MALLS, SAXS and X-ray crystallography, we found no evidence of dimerisation for any allotype, including mutants designed to disrupt previously proposed interfaces^23^. Re-refinement of the original (PDB 3RY6) structural model from which dimerisation had been inferred also eliminated the reported dimer contacts. Crystallisation of the 27W-131H allotype (PDB 8CHA) likewise revealed no dimer interfaces. Together, these findings suggest that earlier reports of FcγRIIa ectodomain homodimers may have arisen from crystal-packing artefacts rather than biologically relevant interactions.

#### Cellular receptor interactions

To address the limitations of isolated ectodomains, we examined full-length FcγRIIa variant behaviour in a membrane environment. Ligand Tracer analysis revealed substantially more complex interaction profiles than those observed by SPR, highlighting the dynamic nature of membrane-bound FcγRIIa. These complex interactions may reflect ligand-driven FcγRIIa clustering and the heterogeneous lateral mobility states previously reported for membrane-bound receptors^54^. A limitation of this approach is the use of monomeric IgG subclasses, which do not fully represent the immune complex-mediated interactions that typically drive FcγRIIa activation *in vivo*. However, employing monomeric ligands enabled direct comparison with the corresponding SPR experiments and facilitated interpretation of allotype-specific binding differences.

While interaction maps revealed IgG subclass binding preferences remained broadly consistent with previous studies^53^, Q27W altered the interaction landscape of IgG3 and disrupted a highly stable interaction component that may correspond to an active signalling state. This finding prompted further functional investigation using signalling reporter assays.

NanoBiT assays demonstrated that both 27Q-131H and 27W-131H receptor allotypes underwent comparable ligand-induced clustering, indicating that Q27W does not substantially impair receptor association. In contrast, recruitment of SYK to activated FcγRIIa was consistently delayed in cells expressing 27W-131H. These results support previous work identifying delayed downstream signalling as the principal functional consequence of the Q27W polymorphism^25^. Furthermore, detection of phosphorylated SYK Tyr-352 confirms that the observed responses represent *bona fide* FcγRIIa-mediated signal transduction^55^.

Taken together, these findings indicate that Q27W has relatively modest effects on IgG binding but can alter the kinetics of FcγRIIa signalling. More broadly, they highlight the importance of studying receptor function within a membrane context, as key signalling-related effects were not apparent from isolated ectodomain analyses alone. These data also suggest that functional consequences of *FCGR2A* polymorphisms may arise through a combination of ligand binding, receptor organisation and downstream signalling properties, potentially further influenced by linked regulatory variants affecting receptor expression.

How delayed signalling contributes to autoimmunity remains uncertain. One possibility is that subtle changes in signalling kinetics alter the balance between activation and regulatory pathways in myeloid cells. Alternatively, delayed signalling could represent a compensatory adaptation that offsets other pro-inflammatory effects of the 2A.3 haplotype. Resolving these possibilities will require investigation in primary immune cells under physiologically relevant stimulation conditions.

#### A model for the biological effects of the 2A.3 haplotype

Our findings suggest that the 2A.3 haplotype exerts both quantitative and qualitative effects on FcγRIIa biology. Quantitatively, it reduces production of soluble FcγRIIa through altered splicing. Qualitatively, it modifies receptor interactions and downstream signalling behaviour. These effects may act in opposing directions, with reduced sFcγRIIa favouring immune activation while altered signalling kinetics potentially dampen aspects of cellular responsiveness. The overall biological consequence of the haplotype is therefore likely to depend on cellular context, immune-complex composition and the relative contribution of different FcγRIIa-expressing cell populations. This complexity may help explain why *FCGR2A* variants have been associated with a broad spectrum of autoimmune and infectious diseases.

## Conclusion

In summary, we identify a common European *FCGR2A* haplotype, 2A.3, as the most likely functional cause of the RA association at this locus. The haplotype combines regulatory effects on sFcγRIIa expression with alterations in receptor signalling behaviour, providing a plausible mechanistic framework linking genetic variation at *FCGR2A* to immune dysregulation. These findings establish a foundation for future studies aimed at understanding how FcγRIIa regulation influences both autoimmunity and host defence.

## METHODS

### Materials and Methods

#### FCGR2A resequencing

A long PCR strategy was adopted to specifically amplify two overlapping long fragments of *FCGR2A*. The first *FCGR2A*-specific long PCR was primed with forward oligo dGTGAGCATTTTAGTACCAGTTGCTTTGAC and reverse dGTGATCATGGCTAGGACTAGAGAGATTCC to amplify a 6.7kb product extending from 8kb upstream incorporating the index SNP rs12746613 to 1.8kb before exon 1. The second fragment was amplified using forward oligo dGTTGGACTGAGGTGGGGTATAGTATATCTCT and reverse dGGAAGCAAAATAAAGGCAATCGGTT to amplify a 15kb product from 2kb upstream of the promoter to the 3’UTR of *FCGR2A*. The target for the forward primer was 8kb from the telomeric end of the segmental duplication in unique sequence and the reverse was in the 3’UTR of *FCGR2A*, which is identical to the *FCGR2C* paralogue. Thus, specificity for *FCGR2A* was conferred by a combination of the unique forward primer target and the lack of amplicon derived from *FCGR2C* due to absence of a corresponding reverse primer binding site. Both long fragments were amplified using Phusion polymerase in HF buffer (New England Biolabs, Hitchin, UK) in a two - step PCR of 32 cycles where the annealing temperature was 72°C and the extension time was 8 minutes, yielding a 15kb product.

Successfully amplified long PCR products were diluted 1:200 with nuclease free water and used as templates in 33 nested PCR reactions which were designed to have minimal overlap spanning the entire long PCR product. Nested PCR products were purified using Invitrogen CST PCR cleanup kits and were sequenced in one direction only using BigDye 3.1 (Thermo Fisher, Waltham, MA, USA). Processed electropherograms were basecalled and aligned to the reference sequence using ClustalW in Bioedit. All acknowledged variants were observed at least twice in the resequencing panel. Pairwise LD (r^2^) was calculated for all confirmed variants using Haploview v4.2 in linkage format ^56^.

In order to distinguish *FCGR2A* SNPs from the homologous *FCGR2B* and *FCGR2C*, we amplified specific long PCR amplicons from each resequencing panel individual using the following oligonucleotide primers: *FCGR2B* forward dCTCCACAGGTTACTCGTTTCTACCTTATCTTAC, reverse dCCCAGAAAGAATCACTTTTAATGTGCTGG (16,660 bp); *FCGR2C* forward dCTCCACAGGTTACTCGTTTCTACCTTATCTTAC, reverse dCCTTTAACAATTCCCCTCTTTTTGTCATCCACTC (20,371 bp), and the same PCR parameters described above for *FCGR2A*. For both *FCGR2B* and *FCGR2C*, minimally overlapping nested PCR amplicons were generated, sequenced and aligned as above. Oligonucleotide primer sequences for the nested PCR products and Sanger sequencing are available on request from the corresponding authors.

### Ethical approvals

All component studies involving human participants were approved by a Research Ethics Committee (REC). The Functional Families (Resequencing) study was approved by the NREC Yorkshire and Humber Leeds East Research Ethics Committee under approval number 04/Q1206/107. The BRAGGSS (Biologics in Rheumatoid Arthritis Genetics and Genomics Study Syndicate) study received ethical approval from the North West Research Ethics Committee under reference COREC 04/Q1403/37. The YEAR (Yorkshire Early Arthritis Register) study was approved by the Yorkshire Research Ethics Committee under approval number MREC/99/3/48. The General RA (RA disease continuum) study received ethical approval from the Leeds West Research Ethics Committee under reference 09/H1307/98. The Sheffield cohort was approved by the South Sheffield Research Ethics Committee under approval number 02/186. The Immunochip study, including WTCCC RA samples, received ethical approval from the North West Research Ethics Committee under reference 99/8/84, while the 1958C control cohort was approved by the North West–Haydock Research Ethics Committee under reference 14/NW/1179. The Spanish rheumatoid arthritis and control sample collection from Granada, Spain was approved by the Ethics Committee of the Spanish Research Council. The UK Biobank, a large-scale prospective epidemiological resource, received ethical approval from the North West Multi-centre Research Ethics Committee under reference 21/NW/0157, with access granted through application 24559. The NIHR BioResource rare diseases RNA phenotyping project (transcriptomics work) was approved by the East of England–Cambridge Central Research Ethics Committee under reference 22/EE/0230.

### Genotyping

DNA samples were available for 1959 White European RA cases and 1120 White European controls. These consisted of: 999 RA cases from the Biologics in Rheumatoid Arthritis Genetics and Genomics Study Syndicate (BRAGGSS) ^57^, 849 RA cases from the Yorkshire Early Arthritis Register (YEAR) ^58^; 111 RA cases from the general rheumatology outpatients in Leeds; 210 controls from Leeds; 910 healthy controls from Sheffield, UK, as previously reported ^59^. Immunochip data were also available for 3869 White European RA cases and 8430 White European controls from the UK and 843 White European RA cases and 1953 White European Controls from Spain that were also genotyped by the Immunochip Consortium ^60^. The Uk cohort included 518 RA cases that were also genotyped using the Sequenom platform.

Genotyping of 1,959 RA cases and 1,120 controls was performed using a Sequenom Mass Array platform according to the manufacturer’s instructions. The Immunochip was an Illumina Infinium High-Density array designed to fine map 186 loci that have been associated with 12 autoimmune diseases ^60^. Quality control (QC) of genotype calling was carried out as described previously^61^. Downstream inclusion criteria were minor allele frequency (MAF) ≥ 0.05, Hardy Weinberg P value (HWP) ≥ 0.001. The post QC Immunochip consortium data were reviewed and 544 markers mapping to the *FCGR* locus and flanking regions (chr1:159,526,365-160,062,520 bp, build 36) were analysed for disease association. Pairwise LD was calculated in healthy controls to check for consistency between cohorts. B allele frequency (BAF) data were extracted from iChip workflow in Illumina Genome Studio. For each SNP assay BAF distribution plots were generated in Excel and manually inspected for shoulders and extra normal distributions.

Sequence verification across the dinucleotide *FCGR2A* Q27W polymorphism was carried out using Sanger (reverse primer) sequencing of an 888 bp PCR product amplified using forward primer dGGAAACAGGATCTTGAGATGGGTCC and reverse dGGGTGGGTGAAATGGGGAATGAATGA.

### Statistical analysis

Exact HWP-values for all Immunochip SNPs available were calculated and plotted against chromosomal position. Each of the 14 SNPs on the Sequenom Plex and 25 SNPs on the Immunochip were analysed for association with RA assuming an additive genetic model using a trend test. This was repeated using a stratified trend test for the 9 SNPs that were common to both datasets. Multivariable logistic regression was used to assess which SNPs were independently associated with disease. Haplotype frequencies and pairwise LD between the CNV and SNPs were calculated in MIDAS ^62^ by treating the copy number as a tri-allelic marker with the three alleles corresponding to deletion, normal and duplication. Double deletions, deletion/duplication combinations and multiple duplications were assumed to be not present. Analysis of haplotype association was performed through the haplotype trend test ^63^.

### Investigation of putative FcγRIIa expression variants

We used the LDhap tool in NCBI LDlink ^70^ to extract haplotype frequencies from populations represented in 1000 genomes, ChIP Atlas to map haplotype SNPs to transcription factor binding sites in *FCGR2A*, and GTEx v8 ^39^ to visualise splice QTLs.

For splice QTL analysis in different blood cell types, we used data from the NIHR BioResource Rare Disease RNA Phenotyping study, which is a multicentre multiomics study of approximately 1,000 patients. It consists of RNA sequencing and proteomics of Platelets, Neutrophils, Monocytes and CD4 T cells, as well as single cell RNA sequencing and ATAC sequencing of PBMC’s with whole-genome sequencing (WGS) for each participant where possible. Whole blood was separated using Lymphoprep with Sepmate (Stem Cell) devices to recover PBMCs, from which monocytes were separated using CD14 positive selection (Stem Cell Human CD14 Positive Selection II 17858). Neutrophils were isolated from whole blood using negative selection (Stem Cell Easysep Human Neutrophil Isolation 19666) and platelets negatively selected from platelet rich plasma using CD45 Dynabeads (Thermo Fisher). RNA was isolated from cell subsets using Trizol Plus (Thermo Fisher) and cDNA libraries generated using the Kapa RNA Hyperprep kit with RiboErase (Roche, Basel, Switzerland) using NovaSeq 6000 and NovaSeq X instruments (Illumina, San Diego, CA, USA).

RNAseq data were processed using the NF-core pipeline (https://github.com/nf-core/rnaseq) incorporating STAR and RSEM. Transcript-level transcripts per million (TPM) values for *FCGR2A* were extracted from merged RSEM transcript TPM files and reshaped using Python (pandas) to generate a sample-by-transcript expression matrix. For each sample, total *FCGR2A* expression was calculated as the sum of TPM values across all 17 transcripts. Expression of the putative soluble *FCGR2A* transcript (ENST00000699277), *FCGR2A* haplotype copy number was derived from genotypes encoding Q27W and H131R from neutrophils. A soluble transcript proportion was calculated as the ratio of soluble transcript TPM to total *FCGR2A* TPM, with division-by-zero values recorded as missing. Data were subsequently reshaped into a participant-level table containing cell type–specific measures for monocyte, neutrophil, and platelet samples. To provide independent evidence of alternative splicing, STAR splice-junction (SJ.out.tab) files were analysed. Read counts supporting exon inclusion and skipping events surrounding exon 5 of *FCGR2A* (chr1:161,513,895– 161,513,932, GRCh38) were extracted and used to calculate percent-spliced-in (PSI) values according to standard cassette-exon methodology. All analyses were performed in a Linux high-performance computing environment using GNU coreutils, awk, Python 3.9, and pandas. Statistical significance was assessed using analysis of variance (ANOVA) applied to linear regression models.

### pQTL analysis in UKBiobank

The UK Biobank genomics and proteomics data sets were used to analyse the effect of *FCGR2A* haplotype copy number on plasma concentration of FcγRIIa. Details on how data were produced are described extensively in Eldjarn *et al*., (2023)^71^. Genotyping for two SNPs defining the haplotypes were available (rs12722986 and rs1801274) for 46,803 individuals for whom the normalised protein expression (NPX) values measured by Olink, were also available. Since only common European haplotypes were analysed, tests were not stratified by ethnicity. Analyses were performed in the UKBB secure computing environment using Kruskal-Wallis H test. Due to high numbers in the analysis groups, effect sizes (η²ᴴ) with 95% confidence intervals were reported only.

### Recombinant FcγRIIa ectodomain protein production

FcγRIIa ectodomain coding sequences were cloned into pOPINTTGneo mammalian expression vector using forward primer d<u>GCGTAGCTGAAACCGGC</u>GCAGCTCCCCCAAAGGCTG and reverse primer d<u>GTGGTGGTGGTGTTT</u>GCTGGGCACTTGGACAGTGATGG, where the underlined portions are complementary to the overhanging acceptor site in the vector. Templates for ligation independent cloning were derived from mRNA isolated from healthy volunteers homozygous for FcγRIIa-131H and 131R. Subsequent variants based on Q27W were generated using Gene Art Site-Directed Mutagenesis PLUS kit (Life Technologies, Warrington, UK). Vector constructs were transiently transfected in to HEK293T cells as described^72^. To purify the secreted His-tagged FcγRIIa ectodomains, a 2-step purification was carried out, first using IgG Sepharose 6 Fast Flow affinity chromatography medium according to the manufacturer’s instructions (GE Healthcare, Buckinghamshire, UK), with a single elution step with 0.5M acetic acid fractionated directly into neutralising 1M Tris HCl pH8.0; secondly pooled fractions were polished using HisTrap Nickel Sepharose (GE Healthcare, Buckinghamshire, UK), eluting in 0.5M NaCl, 0.5M Imidazole. Eluted fractions were pooled and concentrated using Viva Spin 20 devices (GE Healthcare), with 10kDa molecular weight cut-off.

### Surface plasmon resonance

SPR was used to measure affinity constants of FcγRIIa ectodomain allotypes for uncomplexed purified IgG subclasses. In a BIAcore T200, anti-His antibody (GE Life Sciences, Buckinghamshire, England) was amine coupled to all flow cells of an NHS/EDC-activated CM5 sensor chip before deactivating with Ethanolamine, using the manufacturer’s protocol. Purified FcγRIIa ectodomains (1 µg/mL) were captured on flow cells 1-3 by injecting at 5µl/min for 60 s, with flow cell 4 acting as a reference cell. IgG1, IgG2 and IgG3 (The Binding Site, Birmingham, UK) were injected at 30µL/min as analytes from 187.5nM to 3000 nM for 60 s each using single cycle kinetics and PBS-P+ as a running buffer. Sensorgrams were double referenced against flow cell 4. The surface was regenerated using a 60s injection 10 mM Glycine-HCl, pH 1.5, at a flow rate of 30 µL/min. Triplicates were run for each interaction pair.

### Size exclusion chromatography with small angle X ray scattering

To determine the oligomeric states of FcγRIIa 27Q-131H (3.7 mg/mL) and the “S126P” mutant at 2.9 mg/mL, we subjected the purified soluble variants to SEC-SAXS on the B21 SAXS beamline at Diamond Light Source, under proposal SM17069-1. The proteins were first separated on a Superdex 75 5/150 column fitted to an Agilent 1200 HPLC system. Following data collection, *ab initio* envelope modelling was carried out using DAMAVER ^73^ and comparisons between the two proteins compared using SUPCOMB ^68^. Visualisation was carried out in PyMOL Molecular Graphics System, Version 1.2rpre, Schrödinger, LLC).

### Size exclusion chromatography with multi-angle light scattering

We used SEC-MALLS to determine the oligomeric state of purified FcγR ectodomains described above. Proteins were concentrated by centrifugal filtration before being applied to a Superdex 75 5/150 column (Cytiva, Buckinghamshire, UK) with a separation range of 3,000 to 70,000 Mr. The column was flushed for 12 hours with PBS running buffer (0.02% v/v sodium azide (Sigma-Aldrich, Haverhill, UK). Each receptor variant was filtered, then injected onto the column at 0.3 mL/min. Proteins were separated by size and UV absorbance, refractive index and light scattering were measured using a SEC-MALLS QELS system (Wyatt Technology, Haverhill, UK) incorporating a DAWN HELEOS 1184 H28 multi-angle static light scattering detector, a Wyatt QELS+ dynamic light scattering detector and a Wyatt T-rEx 825 differential refractive index detector. Elution profiles were analysed using ASTRA software (Wyatt Technology, Haverhill, UK).

### Structure determination

Protein was buffer exchanged into 75mM NaCl, 5mM Tris-HCl pH 7.4 and concentrated by centrifugal filtration to 8 mg/mL. Crystallisation trials were set up using the JSCG Core Suites I, II, III and IV (Qiagen, Manchester, UK). Crystallisation drops were set up as sitting drops using an NT8 crystallisation robot (Formulatrix, Bedford, MA, USA) mixing 100 nL of protein (100 ng total) in a 1:1 ratio with crystallisation solution from the JSCG Core Suites. Plates were incubated at 20°C and images taken at regular time intervals using a ROCKIMAGER 1000 system (Formulatrix) to monitor crystal formation. At 5 days 12 hours, an ultraviolet two-photon excited fluorescence (UV-TPEF) image was taken to allow discrimination between protein and non-protein crystals.

We also evaluated the published PDB structures 3RY4 (FcγRIIa 27Q-131R) and 3RY5 (FcγRIIa 27Q-131H) for evidence of receptor ectodomain dimerisation using the ClusPro-DC server (cluspro.bu.edu) in dimer mode^69^.

### Ligand Tracer

Full length FcγRIIa was expressed in Sf9 cells for Ligand Tracer experiments. Leucocytes were isolated by density gradient centrifugation using Lymphoprep, according to manufacturer’s recommendations (Stemcell technologies, Vancouver, BC). RNA was extracted using acid phenol RNA method (14), cDNA was generated using Superscript II Reverse transcriptase (Thermo Fisher). In Fusion cloning was used to insert full length FcγRIIa into pFastBac using forward primer: dGACGAGCTCACTAGTCGCGGCCGCATGACTATGGAGACC and reverse: dGGTTTTCCGTACCGGGCTGCAGGTTATTACTGTTGAC. Plasmid variants encoding allotypes 27Q-131R and 27W-131H were introduced using the GeneArt PLUS system (Thermo Fisher) by site directed mutagenesis. Sequence verified pFastBac vectors were transfected into DH10EmBacY competent *E.coli* by heat shock and screened using blue/white selection. Miniprep DNA was prepared from selected white colonies using the PureLink HiPure Plasmid Miniprep kit (Thermo Fisher). Sf9 cells were transfected with Bacmid DNA using X-TremeGENE HP DNA reagent (Roche, Basel, Switzerland). Expression of FcγRIIa was verified using Western blot to measure FcγRIIa expression levels at 24, 48 and 72h time points after the day of proliferation arrest (DPA), with anti-CD32 primary antibody (IV.3), detected with a HRP-conjugated rabbit anti-mouse antibody (P0260), and ECL Plus reagent (Thermo Fisher).

Sf9 cells (DPA+48) expressing full-length FcγRIIa allotypes were captured on a 10cm polystyrene petri dish, pre-treated with biocompatible anchor molecule (SUNBRIGHT® OE-040CS, NOF Corporation, Tokyo, Japan) as described previously ^70^. In a Ligand Tracer Green (Ridgeview Intruments AB, Uppsala, Sweden) fitted with a Red detector, DyLight 650-labelled (Thermo Fisher) IgG1, IgG2 and IgG3 (The Binding Site, Birmingham, UK) were added in two stages to final concentrations of 15nM for 30 mins and 45nM for 90 mins. Sensorgrams were zeroed at baseline, immediately prior to first ligand addition, and normalised to 100%, based on the maximum signal obtained after the 90 min second ligand addition. Sf9 cells, not expressing FcγRIIa were used as a reference for subtraction. Kinetic 1:1 and 1:2 models were fitted to the sensorgrams using the Trace Drawer 1.9.1 software (Ridgeview Instruments AB, Uppsala, Sweden). Interaction maps were generated using the linked option in Trace Drawer (Ridgeview Diagnostics AB, Uppsala, Sweden).

### Heat Aggregated IgG

Heat–aggregated IgG1 (HAG1) was made from therapeutic grade IgG1 CAMPATH-1H ^71^, which was previously stored at −80°C. Heat-aggregated IgG3 (HAG3) was made from IgG3 (The Binding Site, Birmingham, UK) and stored at 4°C. In each case, IgG was heat aggregated by incubating samples at 62°C for 20 minutes. Insoluble material was removed by centrifuging the aliquots at 14000 rpm for 5 minutes at 4°C in a refrigerated Benchtop centrifuge (Eppendorf Corporation, Enfield, CT). HAG1 and HAG3 were frozen in aliquots at −20°C and thawed immediately prior to use.

### Defined Immune Complex production

Defined IgG1 immune complexes were made by hybridising flouroisothiocyanate (FITC)-labelled bovine serum albumin (BSA) with a mouse anti-FITC single chain Fv (scFv) (clone 4M5.3 ^72^) engineered as a human chimeric full IgG1 and expressed in Chinese hamster ovary (CHO) cells followed by protein A affinity purification.

Defined IgG3 immune complexes were made by hybridising oxidised low density lipoprotein (oxLDL) with a mouse anti-oxidised phosphocholine scFv (clone E06 ^73^) engineered as a human chimeric full IgG3 and expressed in CHO cells followed by protein A affinity purification.

### Reporter Cells

HEK293 cells were selected as hosts for stable transfection with vectors expressing FcγRIIa and SYK in the following experiments because they were of human origin with high transfection efficiencies and do not express either FcγRIIa or SYK, nor do they express the monocytic and lymphoblastic marker CD52 which is the antigen for the CAMPATH-1H antibody ^71^. We obtained cells from the ATCC via LGC Standards plc. (Teddington, UK). Cells were grown at 37°C in humidified incubators at 5% CO_2_ in Dulbecco’s modified Eagle’s medium (DMEM) including 10% FCS.

### C terminal LgBiT or SmBiT–tagged FcγRIIa constructs for transient transfection

The enzyme complementation system chosen for analysis of full-length FcγRIIa oligomerisation and exploration of proximal signalling events was the Nanoluc binary technology or nanoBiT (Promega, Madison, WI). This consists of a series of vectors allowing expression of proteins tagged with a split luciferase enzyme whereby the large enzyme fragment (LgBiT) is not functional unless bound to the small fragment (SmBiT) tagged to another molecule. The two parts of the enzyme have a low affinity for each other (190 µM) and the enzyme only becomes active when the tagged molecules are in close physical proximity^74^. We constructed a vector based on pFC34K LgBiT TK neo that expressed full length FcγRIIa tagged with a LgBiT on the C terminal. We also constructed a vector based on pFC36K SmBiT TK neo that expressed FcγRIIa with SmBiT tagged on the C terminal ITAM. The second tyrosine in the ITAM of FcγRIIa is 13 aa from the end of the molecule and there is a 12 aa spacer between the end of the receptor and the enzyme. The sequences encoding full-length FcγRIIa 27Q-131H and 27W-131H allotypes were amplified by PCR from vectors described above for insect cell expression using a High-fidelity DNA polymerase (Phusion, Thermo Fisher). The primers were designed to generate the 954 bp CDS in fragments with 15 bp sequence overlaps with the vector to allow cloning into the Sgf1 – Pme1 digested expression vectors using an In Fusion cloning kit (Takara Bio, Mountain View, CA). The sequences were: forward dGTAAAGCCACCGCTAGCGATCGCCATGACTATGGAGACCCAAATG and reverse dCACCTGAACTGCCTTGGAGCTCGTTATTACTGTTGACATGGTC. The fragments were purified using a Qiagen gel extraction kit (Thermo Fisher) after running the PCRs on a 1% agarose gel. After the In Fusion reaction, competent XL-1 blue bacteria (Stratagene, La Jolla CA) were transformed and put onto Sterilin selective plates (Thermo Fisher) containing 50 µg/mL Kanamycin (Sigma Aldrich, Gillingham, UK). The subsequent colonies were grown in selective LB-Miller medium (10 g/l NaCl) and plasmids harvested using a QIAGEN Spin miniprep kit (QIAGEN, Hilden, Germany). Larger plasmid preparations were made starting with 100 mL cultures of selective LB medium. The plasmids were purified with a QIAGEN midiprep kit and ethanol precipitation. Each FγRIIa allotype was sequence verified and shown to be the transcriptional variant 1 (by comparison to NM_00113629.3). Transfecting both vectors into the same cells allowed studies of spontaneous and immune-complex induced FcγRIIa oligomerisation.

We also generated a SmBiT – SYK fusion – encoding fragments by RT-PCR using THP-1 cells as a source of RNA, using a similar strategy as outlined above. The primer sequences were: forward dGGCTCGAGCGGTGCGATCGCCATGGCCAGCAGCGGCATGGCTG and reverse dGAGCCCGAATTCGTTTAAACTTAGTTCACCACGTCATAGTAGTA. The fragments were cloned into the pFN35K SmBiT TK-neo Flexi vector to produce a N terminal – tagged fusion protein. Sequencing verified the insert was identical to SYK transcript variant 1 (NM_003177.7). When transfected into HEK293 cells along with the vector encoding the FcγRIIa – LgBiT HAIgG binding to the receptor led to a bioluminescent signal when SYK docked with the ITAM on FcγRIIa.

Studies of the kinetics of association of receptors of both allotypes with SYK were carried out using 96 well Nunclon plates (Thermo Fisher). The transient transfections were carried out using the same amounts of plasmid and PEI as described above. Each plate had both QH and WH allotypes represented in nine separate transient transfections. Each transient transfection was added to three wells and the average taken to minimise well-to-well variation. The triplicates were laid out in a checkerboard pattern so that 54 wells were used. Wells at the edge of the plate were not used but were filled with DMEM to reduce evaporation form the active wells. These studies were repeated using equal numbers of stably transfected FcγRIIa and SmBiT-SYK expressing cells plated out in the same pattern. Larger differences were apparent in the cells with higher expression levels.

### Transient transfections

Linear, 25,000Da molecular weight polyethylenimine (PEI) transfection reagent (Alfa Aesar, Heysham, UK) was reconstituted to 1 mg/mL in water heated to 80°C. The pH of the PEI solution was adjusted to pH 7 using concentrated HCl and was subsequently filter sterilised through a Millex-GP 0.22 μM filter (Merck Millipore, Gillingham, UK). PEI was then aliquoted and stored at −80°C. After thawing, tubes were stored at +4°C for a maximum of 2 months before disposal. Aliquots were never refrozen after thawing from −80°C, until use.

Initially, for the quantification of spontaneous dimerisation and studies of FcγRIIa expression by flow cytometry, approximately 200,000 HEK293 cells were transiently transfected in each well of a 6 well plate. Each well received up to 1µg total DNA with 4µg PEI for a 4:1 ratio. Bioluminescence signals were measured using a Berthold plate reader (Berthold Technologies USA, Oak Ridge, TN).

Studies of the kinetics of association of receptors of both allotypes with SYK were carried out using 96 well Nunclon plates (Thermo Fisher). For each triplicate set of wells 300 ng of each plasmid was added in 75 µl of serum – free DMEM. 2400 ng of PEI was added to 75 µl from a 1 mg/mL stock (4:1 ratio of PEI:DNA). The diluted PEI was added to the diluted DNA and the mixture was left at room temperature for 30 minutes for complexes to form. Each transient transfection yielded sufficient cells for three wells, each data point representing the average of three values.

### Bioluminescence

Each plate had both QH and WH allotypes represented in nine separate transient transfections. Each transient transfection was added to three wells and the average taken to minimize well-to-well variation. The triplicates were laid out in a checkerboard pattern so that 54 wells were used. Wells at the edge of the plate were not used but were filled with DMEM to reduce evaporation from the active wells.

Transfected cells were incubated for two days to allow for protein expression. The medium was then removed and replaced with Live Cell Assay Medium containing the proprietary bioluminescent substrate furimazine at a 1:100 dilution (Promega). 50 µl of this medium was added to each well whilst aliquots of diluted heat aggregated IgG were prepared. 50 µl of diluted HA IgG was added to each well to give the desired final concentration of HA IgG. Additions were made at five second intervals so that each well was read after the same length of time for a five second read time.

Each plate was read in a CLARIOstar plate reader (BMG Labtech, Ortenberg, Germany). Averages of three wells were taken as the result of a single transfection. A program of 15 cycles was used which allowed luminescence to be read over the course of approximately one hour.

The average value for each transfection was plotted on graphs of Bioluminescence against time. Similar maximum values were obtained for each allotype. For comparison, each value was normalised so that the QH value was 100%. From each graph the area under the curve was calculated using the midpoint approximation method. A Student’s T test was carried out on the two sets of nine average values from each allotype at each concentration of HA IgG and the P value was calculated.

### Flow Cytometry

In order to demonstrate equal expression from each vector, HEK293 cells were plated out in 6 well plates and transfected with individual FcγRIIa expression vectors containing both allotypes tagged with both LgBiT and SmBiT. After two days incubation at 37°C cells were stained with FITC - labelled anti-CD32 antibodies (AT-10 and IV.3). Both these antibodies have epitopes that map to the IgG binding domain of FcγRIIa: IV.3 binds an epitope which overlaps and blocks the IgG Fc binding domain. Clone IV.3 binds both 131H and 131R allotypes. It is known that AT-10 also recognises the antibody binding domain, therefore the Q27W polymorphism is unlikely to affect the binding of both these antibodies to FcγRIIa. Two antibody clones were used in case of epitope effects conferred by H131R and Q27W polymorphisms. Cells were analysed using a BD LSRII Flow Cytometer (Becton Dickinson, Franklin Lakes, NJ). THP-1 cells were used as a positive control for FcγRIIa expression.

### Stable Cell lines expressing C terminal LgBiT – tagged FcγRIIa and N-terminal SmBiT – tagged SYK

In order to stably express the FcγRIIa allotypes at equal levels the inserts in the transient transfection plasmids described above were shuttled into stable expression vectors. Hence the FcγRIIa – LgBiT vector and SmBiT-SYK vector were used as templates for PCR. The FcγRIIa – LgBiT fragments were generated for both 27Q-131H and 27W-131H allotypes using oligonucleotide primers with homology to the H6Pneo suite of 5 plasmids (1085, 1087, 1088, 1089 and 1090) (LifeArc, Stevenage, UK) and cloned into *Nco*I – *Not*I digested vectors to yield a total of ten vectors. The 1085 vector had no deletions in the promoter whereas the others had increasing numbers of deleted elements leading to lower expression. The SmBiT – SYK fragment was cloned into *Bam*H1 and *Eco*RI digested pFB HYG (Stratagene, La Jolla CA).

8µg of the pFB HYG SmBiT – SYK vector was transfected into a T75 flask of Phoenix A packaging cells (Nolan lab, Stanford University CA) at around 60% confluence. After two days the supernatant (containing replication defective viral particles with a human tropism) was filtered through a 0.45 µm filter (“Minisart” Sartorius AG Gottingen, Germany). The supernatant was used to infect HEK293 cells. The infected HEK293 cells were selected with hygromycin at 500 µg/mL for 10 days.

Once stocks of the cloned SmBiT-SYK expressing cells were created these cells were used as the basis for cells expressing Sm-BiT SYK and FcγRIIa – LgBiT at different expression levels. The H6Pneo vectors described above were transfected into HEK293 cells along with plasmids encoding the gagpol element from a lentiviral vector (pdelta 8.91) and the VSV-G envelope (pMD-VSV-G). After two days the supernatants were filtered as above and added to T75 flasks of HEK293 cells stably expressing SmBiT SYK. The following day the infected cells were selected using G418 containing medium (final concentration 500 µg/mL).

After selection the expression levels were checked using flow cytometry as described above. The 1088 QH and WH vectors expressed FcγRIIa at similar levels to each other. The expression level for each was approximately twice that seen on cells of the undifferentiated THP-1 cell line. Hence the 1088 QH SmBiT – SYK and 1088 WH SmBiT – SYK cells were used for Western blots.

### Western Blots

For Immunoblotting, transient transfections were carried out with vectors described above encoding one of each of the QH and WH allotypes and tagged with LgBiT and one encoding SmBiT-SYK. 1 µg of plasmid was used for each vector for approximately 200,000 HEK293 cells in each well of a 6 well plate. After two days incubation the cells were removed from each well using trypsin and resuspended in 1 mL fresh medium and HA IgG1 was added at 40 µg/mL to activate the receptors. 30 µl of the suspended cells were added to Live Cell Assay buffer containing the substrate for the split enzyme. Bioluminescence was measured and proteins harvested after 10 minutes by centrifugation (500 g, 5 minutes) and the proteins were extracted in RIPA buffer containing “Halt” protease inhibitors (Thermo Fisher) and 1 mM sodium orthovanadate. 100 µg of protein were run out on 12% polyacrylamide gels (BIORAD) and transferred onto PVDF membrane (BIORAD). Blots were blocked in 5% bovine serum albumin (Fraction V Sigma Aldrich, St Louis, MO) before addition of primary antibodies at 1:1000 dilution. Antibodies were washed twice in TBS + 1% Tween20 before addition of the secondary antibody at 1:1000 dilution (P0448 anti rabbit – HRP conjugate, Dako Agilent Technologies, Santa Clara, CA). The following primary antibodies were from Cell Signalling Technologies (Danvers, MA): 2710P rabbit antibody recognising phosphorylated tyrosines at positions 525 and 526; 2701P rabbit antibody against phosphorylated tyrosine 352; 13198S rabbit anti-SYK antibody; 4970 rabbit mAb against β-actin as a loading control.

## Data Availability

All experimental data produced in the present study are available upon reasonable request to the authors. UK Biobank and NIHR BioResource data access is restricted to registered users and is dependent on approvals.

https://www.rcsb.org/structure/8CHA

https://www.internationalgenome.org/data-portal/data-collection/phase3

https://www.ukbiobank.ac.uk/

https://bioresource.nihr.ac.uk/

## Funding

This work was funded jointly by Arthritis UK (grant codes 19764 and 36661) the Medical Research Council (MATURA MR/K015346/1), The Anne Wilks Charitable Foundation, University of Leeds PhD scholarship (EGF), Wellcome Trust Institutional Strategic Support Fund to JIR and NIHR grant numbers RG94028 and RG85445 to the NIHR BioResource RNA Phenotyping Consortium.

We thank all the patients who have contributed to this research, clinical staff who supported patient recruitment and laboratory staff who undertook sample processing. We thank Dr Chi Trinh for protein crystallography support, Amelia Lesiuk for support with Sf9 cell maintenance and baculovirus expression. We thank Dr Felix Randow for gifting the H6Pneo lentiviral vectors incorporated in the reporter assays, which also incorporate original material from Prof Christopher Baum of Hannover Medical School, Germany.

K.L.H. and A.C.B. are supported by the National Institute for Health and Care Research (NIHR) Manchester Biomedical Research Centre. E.W.B., P.E., J.I.R., and A.W.M. are supported in part by NIHR Leeds Biomedical Research Centre. J.D.I. is supported by the NIHR Newcastle Biomedical Research Centre. A.W.M., A.B. and J.D.I. were further supported by NIHR Senior Investigator awards.

This research has been conducted using the UK Biobank Resource under application number 24559. A full list of investigators who contributed to the generation of the data is available from. We thank NIHR BioResource volunteers for their participation, and gratefully acknowledge NIHR BioResource centres, NHS Trusts and staff for their contribution. We thank the National Institute for Health Research and NHS Blood and Transplant. The views expressed are those of the authors and not necessarily those of the NHS, the NIHR or the Department of Health and Social Care.

## Author Contributions

J.I.R., A.W.M., J.H.B. and A.G. designed research; J.I.R., E.W.B., E.G.F., J.C.T., M.T., S.B. and S.E. performed research; S.E.K., S.B., S.E., A.G.W., J.D.I., P.E., J.M., A.B., BRAGGSS and YEAR contributed cohorts, novel reagents or analytic tools; J.I.R., E.W.B., E.G.F., J.C.T., M.T., S.B., A.G., T.B., M.F., J.H.B. and A.W.M. analysed data; J.I.R., E.W.B., E.G.F., J.H.B. and A.W.M. wrote the manuscript.

## Data deposition

Coordinates and structure factors have been deposited in the Protein Data Bank under the accession code 8CHA.

---

**Supplementary Figure 1.**
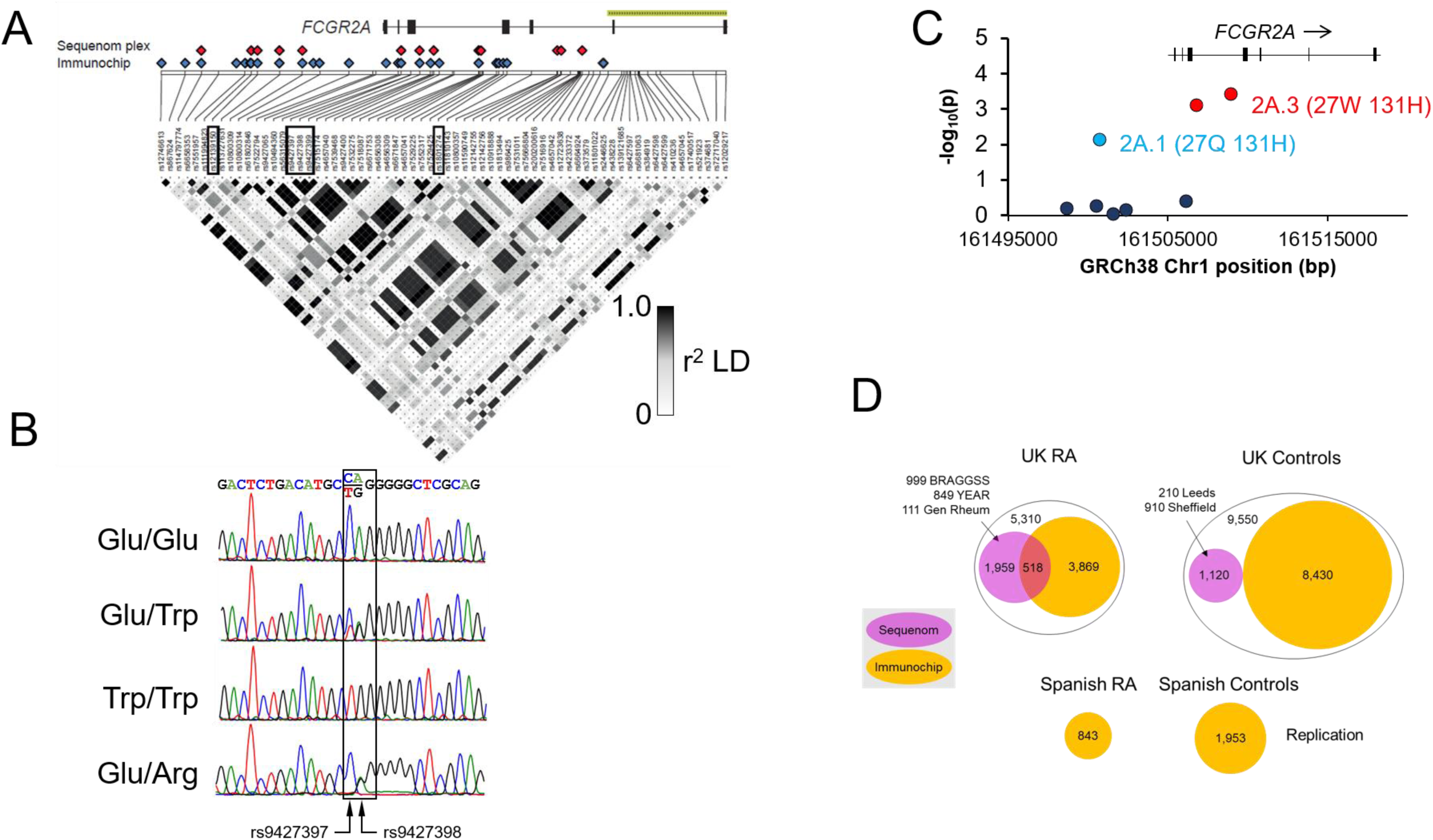
**(A)** Resequencing-confirmed SNPs in *FCGR2A* and pairwise (r^2^) linkage disequilibrium across the segmental duplication boundary in the Caucasian resequencing panel. SNP positions of markers included on custom Sequenom panel and Immunochip marked with red and blue diamonds respectively. Boxed SNPs identify proxy markers utilised for genotyping, where the platform or individual marker assay was confounded by SD. **(B)** Sanger sequencing electropherograms from individuals of different genotypes encoding three alternative amino acids at position 27. **(C)** Close-up of *FCGR2A* for combined iChip and Sequenom genotyping and association p values. Markers coloured by LD (r^2^) with rs12746613, as in Fig. 1(B). **(D)** Association study design. Circle areas are proportional to cohort sizes. BRAGGSS Biologics in Rheumatoid Arthritis Genetics and Genomics Study Syndicate.

**Supplementary Figure 2.**
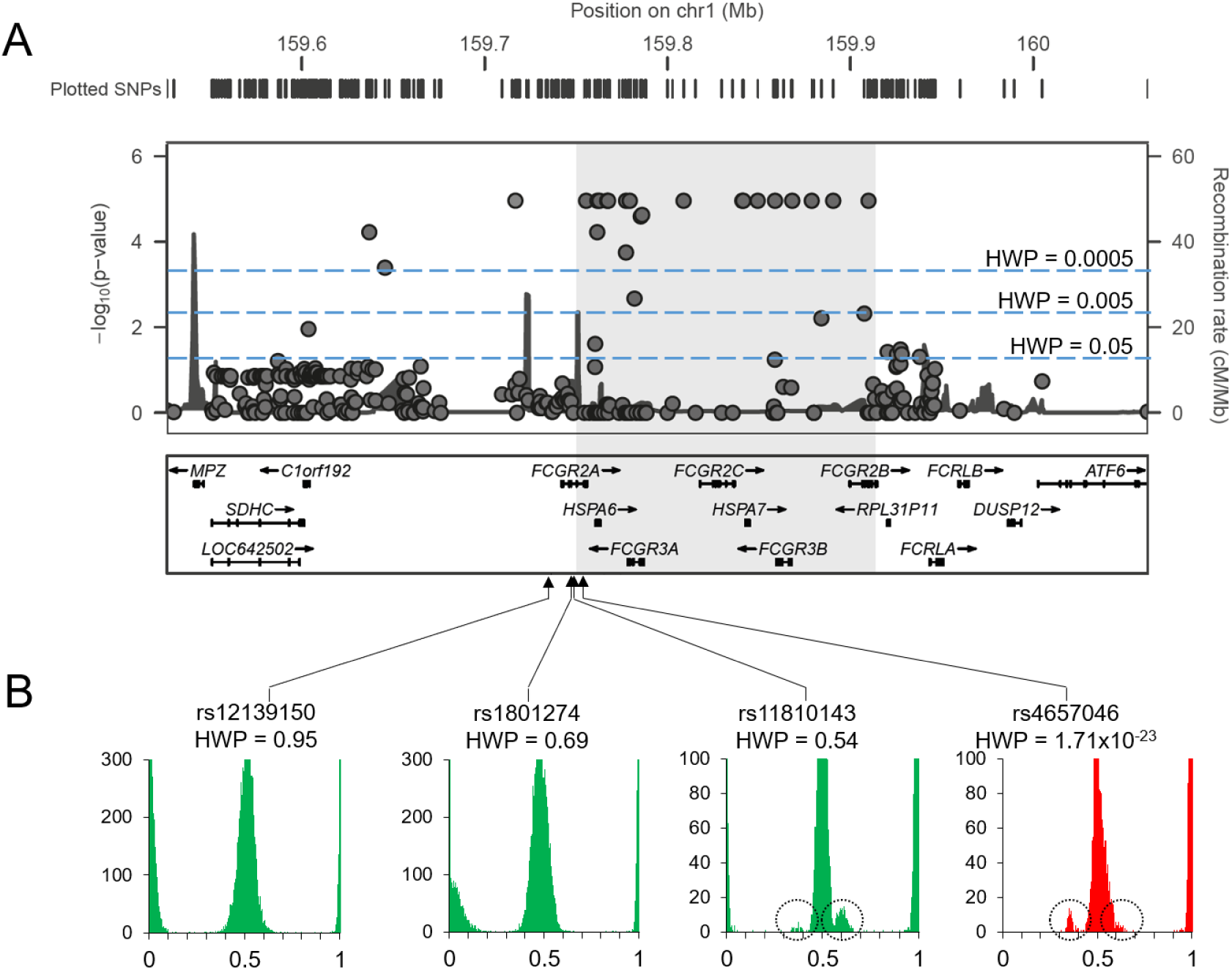
Quality control of 544 Immunochip SNP markers in the segmentally duplicated *FCGR* locus and flanking regions. **(A)** Reported mapping locations of Immunochip SNPs in relation to *FCGR* genes (hg18) and recombination rate. Height on the y axis (negative log of the p-value, capped at 5 for clarity) indicates degree of departure from Hardy Weinberg Equilibrium for the called genotype frequencies. Segmental duplication is indicated by the shaded region. **(B)** B allele frequency (BAF) distribution for selected Immunochip SNP markers inside and outside the *FCGR* segmental duplication. Shoulders and extra peaks suggest CNV in the assay target and/or its paralogs. BAF distributions coloured green for those passing HWP <0.05 criterion, red for fail. HWP Hardy Weinberg p-value.

**Supplementary Figure 3.**
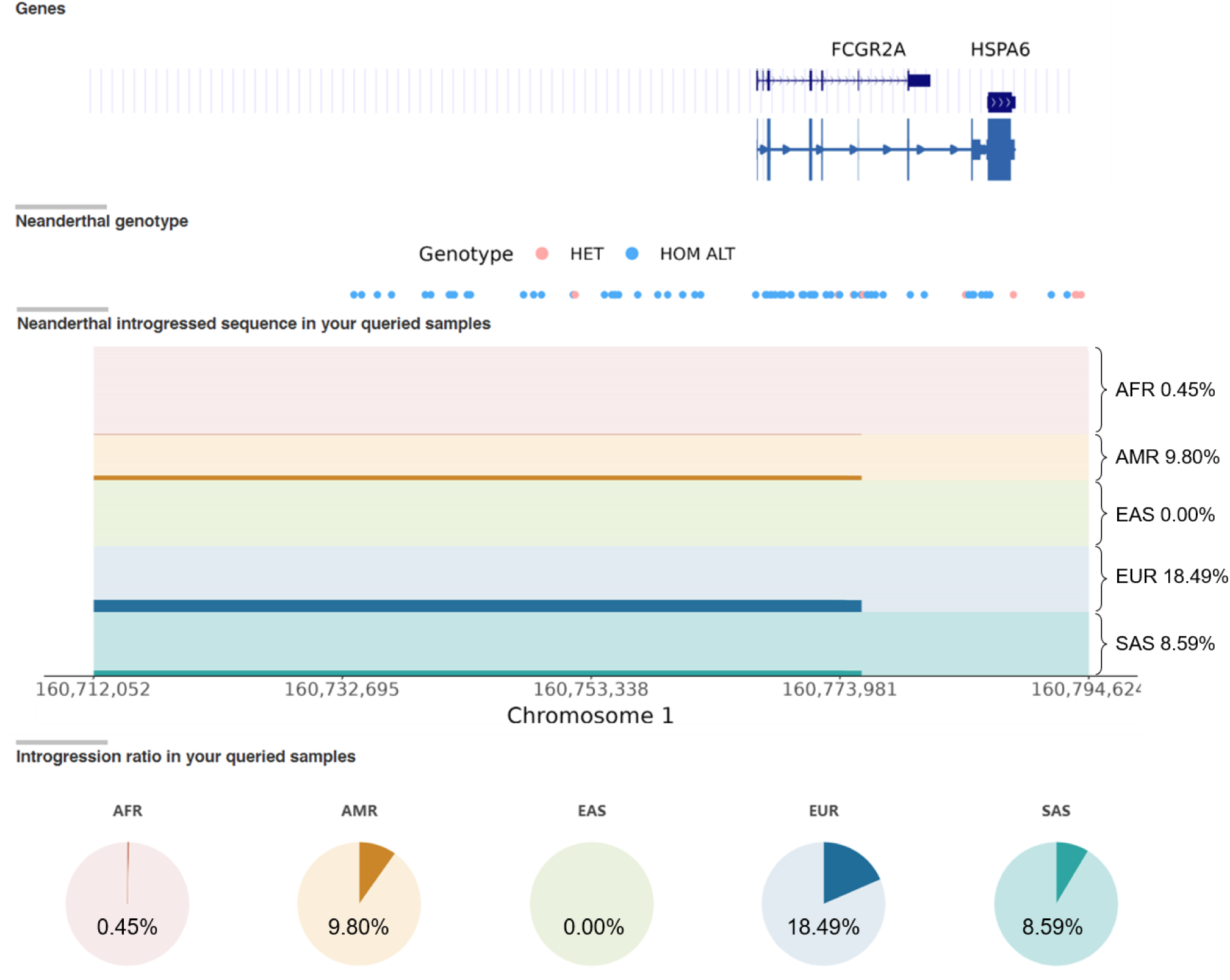
Archaic introgression in *FCGR2A* at chr1:160712052-160794624: output from Arcseqhub using T2T-CHM13v2.0 reference genome. Neanderthal genotypes are shown as red and blue circles along the query genome interval. In introgressed sequence panel, each row is an individual and is organised by population. Altai Neanderthal sequences are plotted in red (African-AFR), orange (Admixed American-AMR), green (East Asian-EAS), dark blue (European-EUR) or shallow blue (South Asian-SAS). The introgression % for all populations are shown in pie charts, respectively.

**Supplementary Figure 4.**
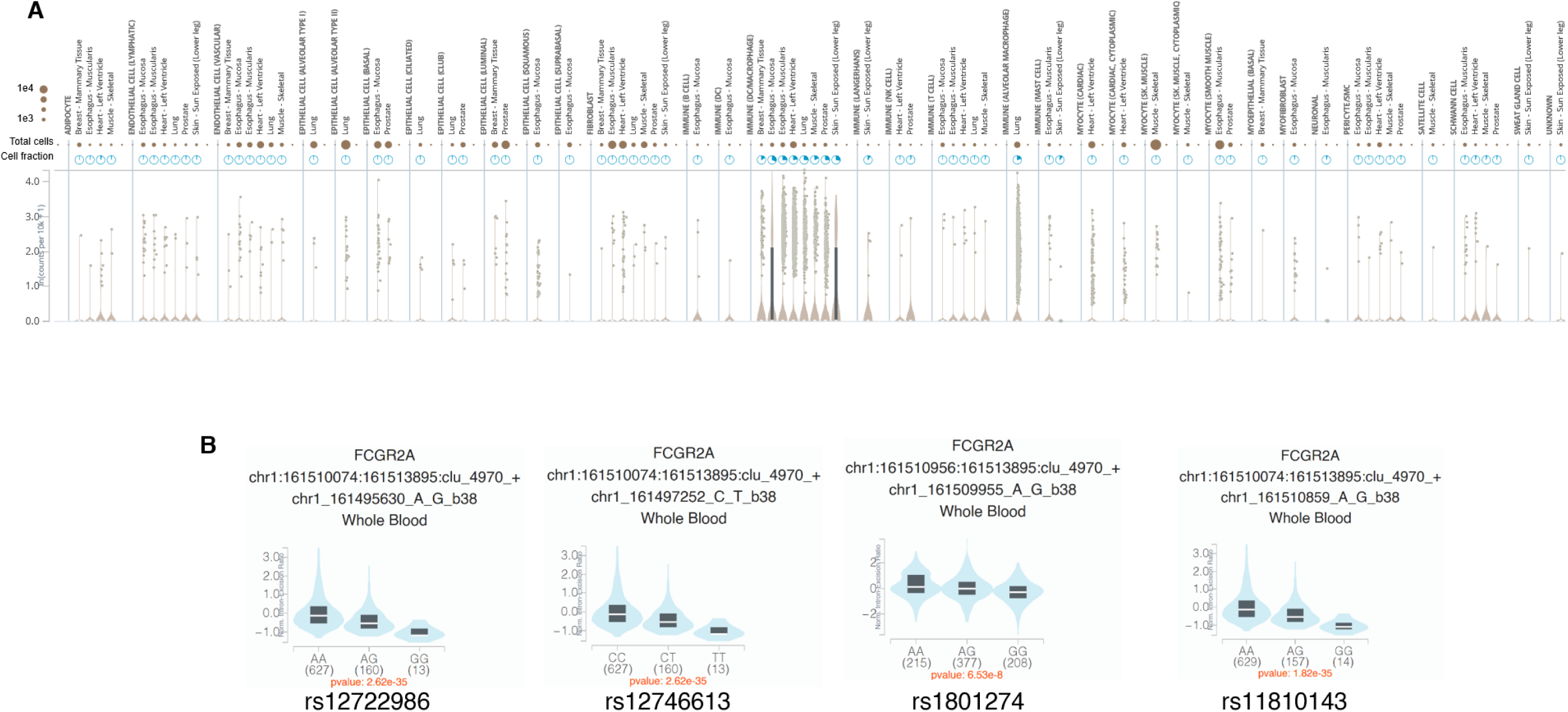
GTEx Analysis Release V10 accessed 13^th^ March 2026. **(A)** Single tissue expression of *FCGR2A* sorted by cell type after single cell RNAseq deconvolution. Gene transcription expressed as ln(counts per 10k +1) to normalise for read differences between tissues. Total cells sequenced per tissue expressed as scaled brown circle areas. Proportion of *FCGR2A-*expressing cells summarised as blue pies. For more details see https://www.gtexportal.org/home/gene/FCGR2A. **(B)** Whole blood splice QTL data for the soluble (missing exon 5) encoding variant of *FCGR2A* for selected SNPs defining the 2A.3 haplotype.

## SUPPLEMENTARY TABLES

**Supplementary Table 1.**
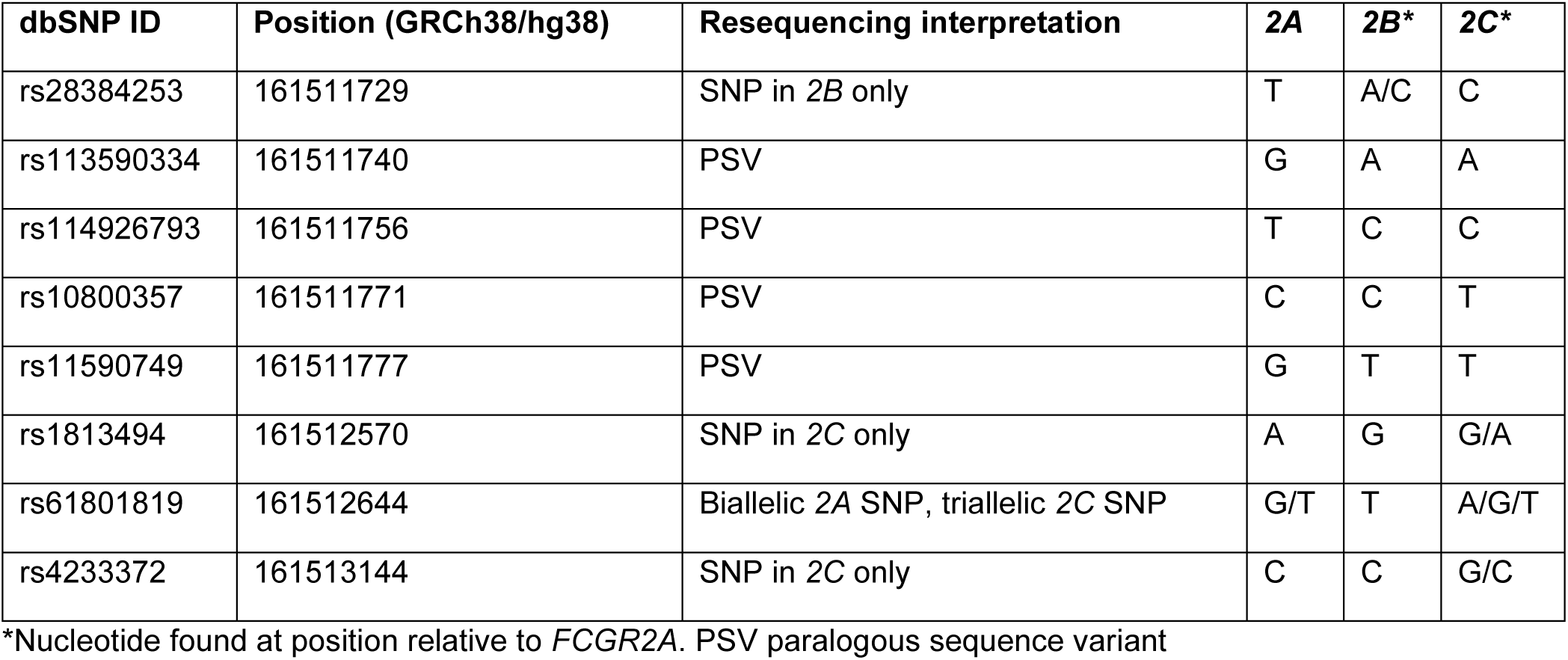
dbSNP (build 155) records mapping to *FCGR2A*, where gene-specific resequencing suggested alternative mapping and variation.

| dbSNP ID | Position (GRCh38/hg38) | Resequencing interpretation | 2A | 2B* | 2C* |
| --- | --- | --- | --- | --- | --- |
| rs28384253 | 161511729 | SNP in 2B only | T | A/C | C |
| rs113590334 | 161511740 | PSV | G | A | A |
| rs114926793 | 161511756 | PSV | T | C | C |
| rs10800357 | 161511771 | PSV | C | C | T |
| rs11590749 | 161511777 | PSV | G | T | T |
| rs1813494 | 161512570 | SNP in 2C only | A | G | G/A |
| rs61801819 | 161512644 | Biallelic 2A SNP, triallelic 2C SNP | G/T | T | A/G/T |
| rs4233372 | 161513144 | SNP in 2C only | C | C | G/C |

\*Nucleotide found at position relative to *FCGR2A*. PSV paralogous sequence variant

**Supplementary Table 2.** Association with rheumatoid arthritis in the Sequenom plex samples (732 cases, 366 controls) for *FCGR2A* Q27W and *FCGR2A* H131R after adjusting for *FCGR3A* F158V and *FCGR3B* deletions.

|  | Unadjusted analysis |  | Multivariable analyses |  |  |  |  |  |  |  |
| --- | --- | --- | --- | --- | --- | --- | --- | --- | --- | --- |
|  | OR | *P* | OR | *P* | OR | *P* | OR | *P* | OR | *P* |
| *FCRG2A* Q27W (proxy rs9427399) | 1.15 | 0.31 | 1.21 | 0.20 | 1.08 | 0.63 |  |  | 1.14 | 0.39 |
| *FCRG2A* H131R (proxy rs12139150) | 1.05 | 0.57 | 1.10 | 0.34 |  |  | 1.10 | 0.31 | 1.13 | 0.22 |
| *FCGR3A* F158V (rs396991) | 1.45 | 0.10 | 1.43 | 0.12 | 1.41 | 1.28 | 1.39 | 0.15 | 1.39 | 0.15 |
| *FCGR3B* deletions | 1.52 | 0.079 |  |  | 1.43 | 1.54 | 1.58 | 0.063 | 1.51 | 0.11 |

**Supplementary Table 3.** Measures of linkage disequilibrium between rheumatoid arthritis associated *FCGR3B* copy number and *FCGR3A* F158V and in healthy controls.

|  |  | CNV *FCGR3B* |  |  | *FCGR3A*-158 |  |
| --- | --- | --- | --- | --- | --- | --- |
|  |  | Deletion<br>(n=50) | Normal<br>(n=412) | Duplication<br>(n=59) | V | F |
| *FCGR2A* | Freq-A (Q) | 0.05 | 0.78 | 0.05 | 0.21 | 0.66 |
| Q27W | Freq-G (W) | 0.00 | 0.12 | 0.00 | 0.12 | 0.02 |
| (rs9427399) | D'(G) | -0.63 | 0.66 | -0.67 | 0.82 | -0.82 |
|  | R<sup>2</sup>(G) | 0.00 | 0.01 | 0.00 | 0.21 | 0.21 |
| *FCGR2A* | Freq-G (R) | 0.03 | 0.46 | 0.04 | 0.09 | 0.43 |
| H131R | Freq-A (H) | 0.02 | 0.44 | 0.01 | 0.23 | 0.25 |
| (rs1801274) | D'(A) | -0.23 | 0.37 | -0.44 | 0.45 | -0.45 |
|  | R<sup>2</sup>(A) | 0.00 | 0.01 | 0.01 | 0.10 | 0.10 |
| *FCGR3A* | Freq-G (V) | 0.03 | 0.27 | 0.01 |  |  |
| F158V |  |  |  |  |  |  |
| (rs396991) | Freq-T (F) | 0.02 | 0.62 | 0.04 |  |  |
|  | D'(V) | 0.33 | -0.08 | -0.21 |  |  |
|  | R<sup>2</sup>(V) | 0.01 | 0.00 | 0.00 |  |  |

**Supplementary Table 4.** Association of UK and Spanish rheumatoid arthritis with the *FCGR2A* Q27W-H131R haplotype *FCGR2A* Q27W was genotyped using rs9427399 as a proxy (27Q-A; 27W-G) and FCGR2A H131R (rs1801274) was genotyped directly (131R-G; 131H-A). NO not observed. NA not applicable.

| Disease Group | Haplotype | Frequency |  | OR | P |
| --- | --- | --- | --- | --- | --- |
|  |  | Controls | Cases |  |  |
| UK RA |  | n=5310? | n=9550? |  |  |
|  | 27Q-131R (2A.2) | 0.53 | 0.51 | 1 |  |
|  | 27W-131R (NO) | 0.00 | 0.00 | NA |  |
|  | 27Q-131H (2A.1) | 0.35 | 0.36 | 1.05 (0.99, 1.12) | 0.087 |
|  | 27W-131H (2A.3) | 0.12 | 0.14 | 1.17 (1.07, 1.27) | **0.0003** |
| Spanish RA |  | n=1953 | n=843 |  |  |
|  | 27Q-131R (2A.2) | 0.511 | 0.508 | 1 |  |
|  | 27W-131R (NO) | 0.000 | 0.000 | NA | NA |
|  | 27Q-131H (2A.1) | 0.359 | 0.332 | 0.93 [0.82-1.06] | 0.261 |
|  | 27W-131H (2A.3) | 0.130 | 0.161 | 1.24 [1.05-1.47] | **0.011** |

**Supplementary Table 5.** Affinity constants of FcγRIIa allotypes for IgG subclasses, measures using surface plasmon resonance and Ligand Tracer. Affinity constants (M) of FcγRIIa allotypes for uncomplexed IgG subclasses measured using surface plasmon resonance and Ligand Tracer experiments. SPR *K_D_* values were determined from steady state responses at concentrations from 187.5nM to 3000nM, in Ligand Tracer *K_D_* values are derived from kinetics of IgG subclass additions to final concentrations of 15nM and 45nM. *For IgG3 and IgG4 on the LT, the interaction was fitted to a 1:2 model, thus two *K_D_* estimates are stated.

|  | IgG1 |  | IgG2 |  | IgG3 |  | IgG4 |  |
| --- | --- | --- | --- | --- | --- | --- | --- | --- |
| FcγRIIa allotype | SPR | LT | SPR | LT | SPR | LT* | SPR | LT* |
| 27Q-131H | 7.9x10<sup>-7</sup> | 4.2x10<sup>-9</sup> | 7.6x10<sup>-7</sup> | 1.4x10<sup>-8</sup> | 3.0x10<sup>-7</sup> | (1) 1.1x10<sup>-7</sup> | n.d. | (1) 5.8x10<sup>-8</sup> |
|  |  |  |  |  |  | (2) 1.6x10<sup>-9</sup> |  | (2) 2.7x10<sup>-9</sup> |
| 27Q-131R | 1.5x10<sup>-6</sup> | 5.0x10<sup>-9</sup> | 2.2x10<sup>-6</sup> | 2.6x10<sup>-8</sup> | 7.9x10<sup>-7</sup> | (1) 3.9x10<sup>-7</sup> | n.d. | (1) 2.1x10<sup>-7</sup> |
|  |  |  |  |  |  | (2) 3.8x10<sup>-10</sup> |  | (2) 4.5x10<sup>-9</sup> |
| 27W-131H | 2.2x10<sup>-6</sup> | 3.5x10<sup>-9</sup> | 1.2x10<sup>-6</sup> | 9.6x10<sup>-9</sup> | 7.9x10<sup>-7</sup> | (1) 5.54x10<sup>-7</sup> | n.d. | (1) 2.2x10<sup>-7</sup> |
|  |  |  |  |  |  | (2) 3.55x10<sup>-9</sup> |  | (2) 2.2x10<sup>-9</sup> |

**Supplementary Table 6.** Data collection and refinement statistics for FcγRIIa 27W ectodomain (PDB 8CHA)

|  |  |
| --- | --- |
| **Data collection temperature (K)** | 100 |
| **Diffraction source** | Diamond Synchrotron I04 |
| **Wavelength (Å)** | 0.9795 |
| **Detector** | Dectris Pilatus 2M pixel |
| **Refinement** | Phenix v1.13 |
| **Model building** | Coot |
| **Data scaling** | Aimless |
| **Phasing** | Phaser |
| **Structure solution method** | Molecular replacement with 3RY4 |
| **Resolution range (Å)** | 41.82 - 1.81 (1.875 - 1.81) |
| **Space group** | C 2 2 2<sub>1</sub> |
| **Unit cell (Å, °)** | 49.9, 76.5, 108.3, 90, 90, 90 |
| **Unique reflections** | 19293 (1867) |
| **Multiplicity** | 6.1 (4.0) |
| **Completeness (%)** | 99.00 (96.8) |
| **Mean I/σ(I)** | 6.3 (1.4) |
| **Wilson B-factor (Å<sup>2</sup>)** | 21.73 |
| **R-merge** | 0.15 (0.73) |
| **R-meas** | 0.16 (0.83) |
| **R-pim** | 0.06 (0.39) |
| **CC<sub>1/2</sub>** | 0.99 (0.45) |
| **Reflections used in refinement** | 19122 (1828) |
| **Reflections used for R-free** | 907 (94) |
| **R-work** | 0.193 (0.361) |
| **R-free** | 0.225 (0.344) |
| **Number of non-hydrogen atoms** | 1587 |
| **macromolecules** | 1349 |
| **ligands** | 127 |
| **solvent** | 111 |
| **Protein residues** | 168 |
| **RMS (bonds)** | 0.004 |
| **RMS (angles)** | 0.74 |
| **Ramachandran favored (%)** | 98.78 |
| **Ramachandran allowed (%)** | 1.22 |
| **Ramachandran outliers (%)** | 0.00 |
| **Rotamer outliers (%)** | 0.00 |
| **Clashscore** | 1.40 |
| **Average B-factor** | 32.17 |
| **macromolecules (Å<sup>2</sup>)** | 29.48 |
| **ligands (Å<sup>2</sup>)** | 55.55 |
| **solvent (Å<sup>2</sup>)** | 38.10 |
| **Number of TLS groups** | 6 |

1217 See PDB validation report for additional structure factors

